# High-grade B-cell lymphoma, not otherwise specified: an LLMPP study

**DOI:** 10.1101/2025.03.11.25323696

**Authors:** Brett Collinge, Laura K. Hilton, Jasper Wong, Waleed Alduaij, Susana Ben-Neriah, Graham W. Slack, Pedro Farinha, Merrill Boyle, Barbara Meissner, James R. Cook, German Ott, Andreas Rosenwald, Elias Campo, Catalina Amador, Timothy C. Greiner, Philipp W. Raess, Joo Y. Song, Giorgio Inghirami, Sarah L. Ondrejka, Elaine S. Jaffe, Dennis D. Weisenburger, Wing C. Chan, Harald Holte, Klaus Beiske, Kai Fu, Jan Delabie, Stefania Pittaluga, Javeed Iqbal, George Wright, Kerry J. Savage, Andrew J. Mungall, Louis M. Staudt, Christian Steidl, Andrew L. Feldman, Ryan D. Morin, Lisa M. Rimsza, David W. Scott

## Abstract

Molecular characterization of high-grade B-cell lymphoma, not otherwise specified (HGBCL-NOS), is hindered by its rarity, evolving definition, and poor diagnostic reproducibility. To address this challenge, we analyzed 92 HGBCL-NOS tumors collected across Lymphoma/Leukemia Molecular Profiling Project sites. Leveraging comparison cohorts of diffuse large B-cell lymphoma (DLBCL-NOS) and Burkitt lymphoma (BL), and molecular frameworks described in these entities, our analysis revealed a heterogenous molecular landscape, reminiscent of DLBCL-NOS but with an enrichment of BL features. By cell-of-origin, 59% were germinal center B-cell-like (GCB), and 25% were activated B-cell-like (ABC). LymphGen, a genetic classifier for DLBCL-NOS, assigned a genetic subtype to 34% of HGBCL-NOS. Although classification rate was lower than in DLBCL-NOS (66%), assigned subtypes spanned the spectrum of LymphGen classes, including 31% of ABCs classified as MCD. Features differentiating HGBCL-NOS from DLBCL-NOS included *MYC*-rearrangement (47% vs. 6%), dark zone signature (DZsig) expression (45% vs. 7%), and more frequent mutation of *ID3*, *MYC*, *CCND3*, and *TP53* – all common to BL. A genetic classifier that differentiates DLBCL-NOS from BL classified 53% of DZsig+ tumors as BL-like, with those classified as DLBCL-like frequently *BCL2*-rearranged. Among DZsig-GCB tumors, 95% were DLBCL-like. Centralized pathology review reclassified almost half of tumors as DLBCL-NOS but did not identify a more homogenous HGBCL-NOS population, with no difference in features between confirmed and reclassified tumors. In conclusion, molecular testing enables a subset of HGBCL-NOS to be assigned to established categories. Based on rarity and diagnostic challenges, broader inclusion of HGBCL-NOS should be considered in biomarker-driven DLBCL trials.

**Key points:**

1. Molecular analyses reveal that HGBCL-NOS encompasses a heterogeneous collection of tumors.
2. A subset of HGBCL-NOS can be assigned to established molecular groups, while others remain unclassified.

## Introduction

High-grade B-cell lymphoma, not otherwise specified (HGBCL-NOS), is a diagnostic entity in which membership is defined by malignant cell morphology (i.e. blastoid chromatin resembling lymphoblastic lymphoma or cytologic features intermediate between Burkitt lymphoma [BL] and diffuse large B-cell lymphoma [DLBCL]), and the absence of concurrent *MYC-* and *BCL2-* and/or *BCL6-*rearrangements.^1–4^ This group represents an evolving diagnostic category formally introduced in the 1994 REAL classification when it was recognized that some tumors could not be confidently classified as BL or DLBCL, giving rise to the provisional entity “High-grade B-cell lymphoma, Burkitt-like”.^5^ The most recent refinement in the evolution of this category has been the establishment^1^, and subsequent refinement, of high-grade B-cell lymphoma/DLBCL with *MYC* and *BCL2*-rearrangements (HGBCL-DH-*BCL2*), noting that the latest classifications are at variance with tumors with *MYC-* and *BCL6-*rearrangements^2,3,6^. Other efforts to refine HGBCL-NOS have primarily focused on restricting its classification by more clearly defining BL and DLBCL, resulting in minimal material changes to the overall classification of this group^7,8^. Consequently, HGBCL-NOS remains a diagnosis of exclusion that largely relies on morphological assessment.

A recent large retrospective multi-institutional study of HGBCL-NOS highlights the unmet needs and challenges of this group.^9^ Most patients had high-risk features, including stage IV disease and elevated lactate dehydrogenase. Reflecting the lack of standard treatment for this group, patients were treated with a range of regimens, from the DLBCL standard (R-CHOP) to more intensive regimens typically reserved for BL and HGBCL-DH-*BCL2*. However, outcomes were not improved with intensified treatment, with progression-free survival (PFS) of 55% at 2 years, emphasizing the need for more effective treatment strategies. Characterization of the tumors by immunohistochemistry (IHC) and fluorescence *in situ* hybridization (FISH) revealed considerable heterogeneity beyond the majority having a germinal center B-cell-like (GCB) immunophenotype. This heterogeneity has also been observed in other studies that employed targeted sequencing approaches.^10–14^

Optimal classification identifies tumors with common biological, clinical, and molecular features, while maintaining accurate tumor categorization and minimizing the number of unclassified cases. BL and DLBCL-NOS have been extensively characterized at the genomic level, revealing their core genetic aberrations and underlying molecular substructure.^15–20^ In contrast, HGBCL-NOS remains poorly understood, and improving its clinical management requires a better understanding of this group. However, progress has been hindered by its rarity and the challenges associated with membership being contingent on identifying high-grade morphology, which is affected by pre-analytic variables and inter-observer variability. To overcome these barriers, we collected HGBCL-NOS tumors from affiliated sites of the Lymphoma/Leukemia Molecular Profiling Project (LLMPP) consortium. Leveraging the framework provided by our understanding of the BL and DLBCL, we conducted a comprehensive molecular analysis of HGBCL-NOS to determine whether these tumors share common molecular features or if they can be more accurately classified using molecular approaches. To reflect the real-world setting, analyses were initially performed on tumors based on their submitted local diagnosis. All tumors further underwent centralized pathology review by the LLMPP pathology panel and were reanalyzed to assess whether this process could identify a more cohesive group.

## Methods

### Study cohort

Ninety-two patients with an available HGBCL-NOS biopsy were identified from eight LLMPP affiliated sites (supplemental Table 1). All biopsies were locally reviewed by an LLMPP hematopathologist to confirm a HGBCL-NOS diagnosis and to assign a cytological subtype following WHO revised 4^th^ edition criteria^21^. FISH was performed on all biopsies to exclude HGBCL-DH-*BCL2* and HGBCL-DH-*BCL6*. A relapse biopsy was included for seven patients for where the primary biopsy was unavailable. Five patients had a known prior indolent lymphoma. Patients were excluded from the study if they were <18 years of age or had known HIV infection.

### FISH and IHC

Tissue microarrays (TMAs) were constructed from duplicate 0.6mm cores. FISH was performed on the TMAs using break-apart probes for *MYC*, *BCL2,* and *BCL6*. IHC was performed on TMA sections using commercially available antibodies and independently scored by two pathologists. IHC positivity was defined as ≥30% (CD10, BCL6, and MUM1)^22^, ≥40% (MYC), and ≥50% (BCL2)^23^ staining of tumor cells. For biopsies not included on TMAs, clinical IHC and FISH data provided by the submitting institution were used when available.

### Detection of somatic mutations, copy number alterations, and structural variants

Whole genome sequencing (WGS, n = 12) or whole exome sequencing (WES, n = 49) was performed for 61 samples. Biopsies with available fresh frozen tissue underwent WGS, while WES was performed on formalin-fixed paraffin-embedded (FFPE) tissue. Matched normal samples were available for six cases. Tissue pairing and preservation status, as well as coverage and quality metrics, are included in supplemental Table 1. Somatic mutations were identified with a consensus variant calling approach, SLMS-3, using Strelka2^24^, LoFreq^25^, SAGE, and Mutect2^26^, as previously described^27^ (supplemental Table 2). Significantly mutated genes were identified using dNdScv^28^, MutSig2CV^29^, and OncodriveFML^30^. LymphGen classification^17^ was performed on variant calls using somatic mutations and *BCL2/BCL6-*rearrangement data. Variant calls were further used to assign tumors into BL-like and DLBCL-like genetic subtypes using a previously described random forest classifier.^20^ Structural variants involving *MYC* and *BCL2* were called using Manta^31^ and GRIDSS2^32^ on WGS (n = 12) or hybridization capture sequencing (n = 54) using a panel designed to capture the regions around *MYC*, *BCL2*, and recurrent rearrangement partner loci, as previously described^33^ (supplemental Table 3). Copy number alterations (CNAs) were identified from WGS (n = 12) or low pass WGS (n = 46) using Control-FREEC^34^ (supplemental Table 4), and recurrent CNAs were identified using GISTIC2^35^.

### Gene expression profiling

RNA sequencing was performed on RNA extracted from FFPE material for 58 biopsies using a ribodepletion-based protocol. Gene expression was quantified using Salmon^36^ , and differential gene expression analysis was performed using DEseq2^37^. MiXCR^38^ was used to predict full-length immunoglobulin transcripts of the dominant clone. Cell-of-origin (COO) and dark zone signature (DZsig) status were determined using the DLBCL90 assay on the nCounter platform (nanoString), as previously described.^39^ Tumors were assigned to refined COO subgroups, prioritizing COO over DZsig status for activated B-cell-like (ABC) tumors. DZsig+ GCB and unclassified tumors were grouped as DZsig+, while DZsig- and DZsig-indeterminant tumors remained in their respective COO subgroups.^40^

### Comparison of molecular features in HGBCL-NOS to BL and DLBCL

Data from two previously described cohorts were used to compare the molecular features of HGBCL-NOS with BL and DLBCL. The first cohort included 63 BL biopsies with WGS data from the Burkitt Lymphoma Genome Sequencing Project (BLGSP).^20^ This cohort was limited to patients aged 18 or older with EBV-negative tumors (supplemental Tables 5-6) to align with the HGBCL-NOS cohort inclusion criteria and avoid EBV status as a confounding factor, as EBV-positive BL has distinct molecular features^19,20^. The second cohort consisted of 781 DLBCL-NOS biopsies, drawn from a population-based cohort of adult patients diagnosed with a *de novo* tumor of DLBCL morphology between 2005 and 2010 in British Columbia, Canada (supplemental Table 7).^40^ Data available for this cohort included FISH, IHC, and DLBCL90 gene expression profiling. All biopsies had sufficient FISH data to exclude a diagnosis of HGBCL-DH. Targeted capture sequencing of 125 genes that are recurrently mutated in aggressive B-cell lymphomas was performed on 341 FFPE biopsies with adequate nucleic acids (supplemental Tables 8-9). Coverage and quality metrics are provided in supplemental Table 7. A summary of all sequencing data included in this study is available in supplemental Table 10.

### Centralized Pathology Review

All tumors included in this study underwent subsequent centralized pathology review (CPR), conducted by a panel of LLMPP hematopathologists. Each tumor was independently assessed by four pathologists, and a diagnosis was finalized if at least three agreed. If fewer than three agreed, the case was reviewed by the entire panel (10-15 pathologists) during an in-person or virtual meeting, with the final diagnosis determined by majority vote. Analyses are interpreted according to the initial local submitting diagnosis unless otherwise stated.

### Statistical Analysis and Ethics Approval

Statistical analyses were performed with R version 4.4.0. This study was approved by the University of British Columbia/BC Cancer Research Ethics Board in accordance with the Declaration of Helsinki. All patient and sample identifiers have been replaced with research-specific identifiers.

## Results

### Clinical and molecular characteristics of the study cohort

This study included 92 biopsies submitted as HGBCL-NOS from eight LLMPP sites (supplemental Table 1). For comparisons of HGBCL-NOS with other related aggressive B-cell lymphoma entities, 63 BL and 781 DLBCL-NOS biopsies were drawn from previously described cohorts^20,40^ and included in analyses when indicated (supplemental Figure 1). HGBCL-NOS tumors in the main study cohort were identified by the submitting pathologists as showing the following morphologic features: 55% with features intermediate between BL and DLBCL (henceforth referred to as “intermediate” morphology), 34% blastoid cytology, and 11% unspecified (Figure 1A). Clinical characteristics of the study cohort are included in Table 1. There was no significant difference between the clinical characteristics of blastoid and intermediate tumors.

**Figure 1.**
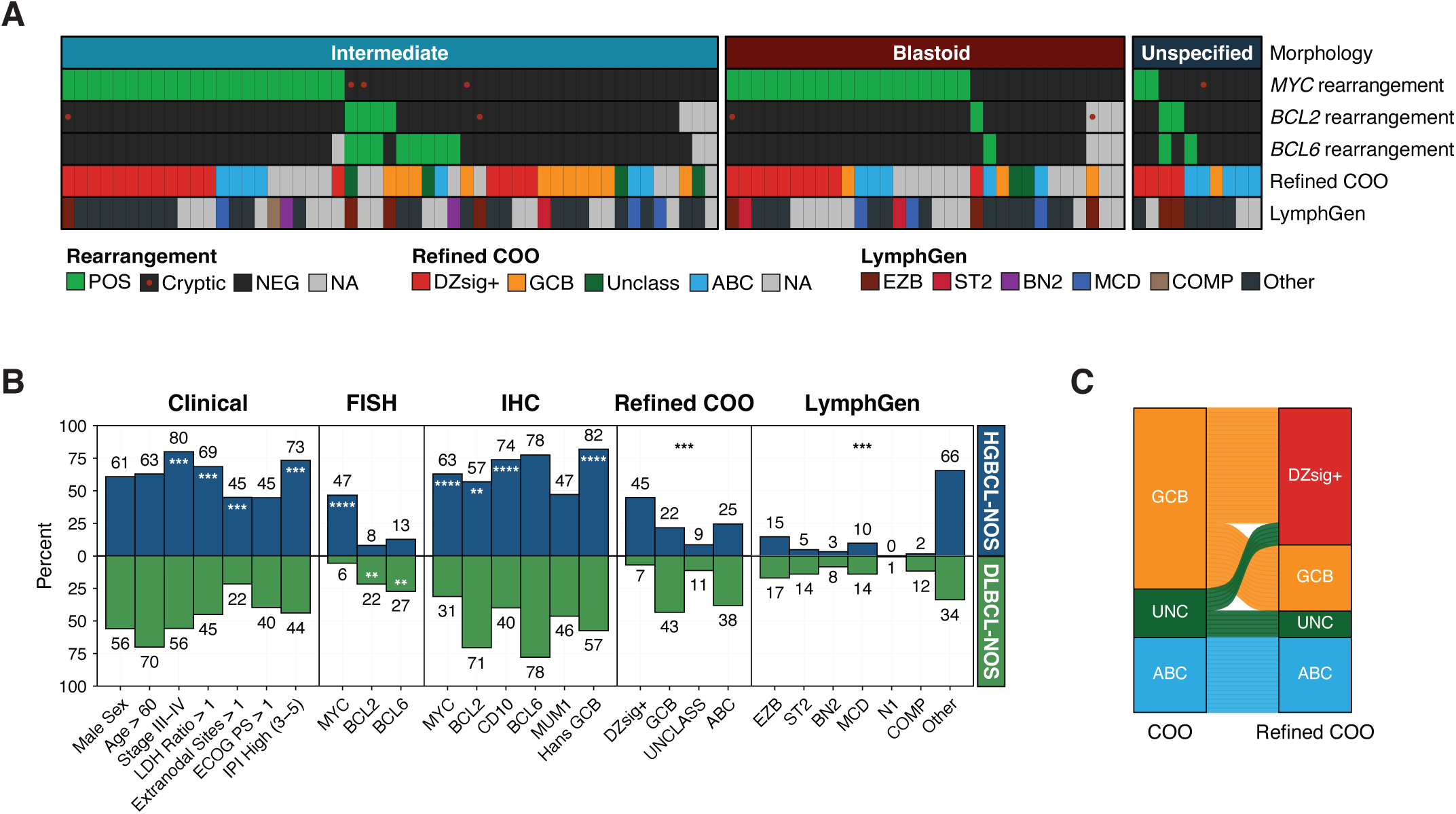
Clinical and molecular features of HGBCL-NOS. **(A)** Heatmap of molecular features of the 92 HGBCL-NOS tumors included in the study, grouped by morphological subtype. *MYC*, *BCL2*, and *BCL6* rearrangement status were determined by FISH. Tumors with a negative (or missing) *MYC* or *BCL2* FISH result where a rearrangement was identified by sequencing are indicated with a red dot. **(B)** Comparison of clinical and molecular features in HGBCL-NOS to DLBCL-NOS. P values were calculated by Fisher’s exact test for comparisons of clinical, FISH, and IHC data, and by χ2 test for Refined COO and LymphGen comparisons. **(C)** Distribution of COO and refined COO subtypes in HGBCL-NOS. **\***P < 0.05; **\*\***P < 0.01; **\*\*\***P < 0.001; **\*\*\*\***P < 0.0001.

**Table 1:** Clinical and pathological characteristics of tumors submitted as HGBCL-NOS by local pathology review. * Comparison between intermediate and blastoid morphology groups LDH: lactate dehydrogenase; ULN: upper limit of normal; ECOG PS: Eastern Cooperative Oncology Group Performance Status; IPI: International Prognostic Index; NA: not available; GCB: germinal center B-cell-like; ISH: *in situ* hybridization; COO: cell-of-origin; ABC: activated B-cell-like; FISH: fluorescence *in situ* hybridization.

| Characteristic | Total cohort | Intermediate morphology | Blastoid morphology | Unspecified morphology | P value* |
| --- | --- | --- | --- | --- | --- |
|  | N = 92 | n = 51 | n = 31 | n = 10 |  |
| Age > 60, n (%) | 58 (63) | 33 (65) | 17 (55) | 8 (80) | 0.49 |
| Age, median (range) | 66.5 (19 – 90) | 66 (30 – 90) | 66 (22 – 89) | 73.5 (19 – 84) | 0.48 |
| Male sex, n (%) | 56 (61) | 32 (63) | 21 (68) | 3 (30) | 0.81 |
| Ann Arbor stage, n (%) |  |  |  |  |  |
| I/II | 10 (20) | 7 (25) | 3 (16) | 0 (0) | 0.72 |
| III/IV | 40 (80) | 21 (75) | 16 (84) | 3 (100) |  |
| missing | 42 | 23 | 12 | 7 |  |
| LDH > ULN, n (%) |  |  |  |  |  |
| No | 16 (31) | 11 (38) | 3 (17) | 2 (50) | 0.19 |
| Yes | 35 (69) | 18 (62) | 15 (83) | 2 (50) |  |
| missing | 41 | 22 | 13 | 6 |  |
| Extranodal sites, n (%) |  |  |  |  |  |
| 0-1 | 33 (55) | 21 (57) | 12 (52) | - | 0.79 |
| >1 | 27 (45) | 16 (43) | 11 (48) | - |  |
| missing | 32 | 14 | 8 | 10 |  |
| ECOG PS, n (%) |  |  |  |  |  |
| 0-1 | 26 (55) | 16 (62) | 9 (50) | 1 (33) | 0.54 |
| 2-4 | 21 (45) | 10 (38) | 9 (50) | 2 (67) |  |
| missing | 45 | 25 | 13 | 7 |  |
| IPI, n (%) |  |  |  |  |  |
| Low (0 – 2) | 12 (27) | 9 (33) | 3 (18) | - | 0.32 |
| High (3 – 5) | 33 (73) | 18 (67) | 14 (82) | 1 (100) |  |
| missing | 47 | 24 | 14 | 9 |  |
| Transformed, n (%) |  |  |  |  |  |
| No | 53 (91) | 29 (97) | 18 (86) | 6 (86) | 0.29 |
| Yes | 5 (9) | 1 (3) | 3 (14) | 1 (14) |  |
| unknown | 34 | 21 | 10 | 3 |  |
| Relapse biopsy, n (%) |  |  |  |  |  |
| No | 53 (90) | 26 (84) | 20 (95) | 7 (100) | 0.38 |
| Yes | 6 (10) | 5 (16) | 1 (5) | - |  |
| unknown | 33 | 20 | 10 | 3 |  |
| Immunohistochemistry |  |  |  |  |  |
| CD10 | 65/88 (74%) | 37/48 (77%) | 24/31 (77%) | 4/9 (44%) | 1 |
| BCL6 | 55/71 (77%) | 33/41 (80%) | 14/21 (67%) | 8/9 (89%) | 0.35 |
| MUM1 | 25/53 (48%) | 13/30 (43%) | 8/14 (57%) | 4/9 (44%) | 0.52 |
| MYC | 34/54 (63%) | 20/34 (59%) | 10/14 (71%) | 4/6 (67%) | 0.52 |
| BCL2 | 49/86 (57%) | 25/50 (50%) | 16/27 (59%) | 8/9 (89%) | 0.48 |
| TDT | 1/49 (2%) | 0/25 (0%) | 1/15 (7%) | 0/9 (0%) | 0.38 |
| Hans COO, n (%) |  |  |  |  |  |
| GCB | 68 (82) | 39 (87) | 25 (86) | 4 (44) | 1 |
| Non-GCB | 15 (18) | 6 (13) | 4 (14) | 5 (56) |  |
| missing | 9 | 6 | 2 | 1 |  |
| Ki67, median (range) | 90 (40 – 100) | 90 (40 – 100) | 100 (70 – 100) | 90 (50 – 100) | 0.35 |
| EBER ISH positive | 1/49 (2%) | 0/33 (0%) | 1/13 (8%) | 0/3 (0%) | 0.28 |
| COO, n (%) |  |  |  |  |  |
| ABC | 17 (25) | 7 (18) | 5 (25) | 5 (50) | 0.75 |
| Unclassified | 11 (16) | 7 (18) | 4 (20) | - |  |
| GCB | 41 (59) | 25 (64) | 11 (55) | 5 (50) |  |
| missing | 23 | 12 | 11 | - |  |
| Refined COO, n (%) |  |  |  |  |  |
| ABC | 17 (25) | 7 (18) | 5 (25) | 5 (50) | 0.69 |
| Unclassified | 6 (9) | 4 (10) | 2 (10) | - |  |
| GCB | 15 (22) | 11 (28) | 3 (15) | 1 (10) |  |
| DZsig+ | 31 (45) | 17 (44) | 10 (50) | 4 (40) |  |
| missing | 23 | 12 | 11 | - |  |
| Rearrangement (FISH) |  |  |  |  |  |
| *MYC* | 43/92 (47%) | 22/51 (43%) | 19/31 (61%) | 2/10 (20%) | 0.17 |
| *BCL2* | 7/86 (8%) | 4/48 (8%) | 1/28 (4%) | 2/10 (20%) | 0.65 |
| *BCL6* | 11/86 (13%) | 8/48 (17%) | 1/28 (4%) | 2/10 (20%) | 0.14 |
| Cryptic HGBCL-DH-*BCL2* | 4/66 (6%) | 3/39 (8%) | 1/19 (5%) | 0/8 (0%) | 1 |
| LymphGen, n (%) |  |  |  |  |  |
| EZB | 9 (15) | 4 (12) | 3 (15) | 2 (29) | 0.69 |
| ST2 | 3 (5) | 1 (3) | 2 (10) | - |  |
| BN2 | 2 (3) | 2 (6) | - | - |  |
| MCD | 6 (10) | 3 (9) | 3 (15) | - |  |
| Other | 40 (66) | 23 (68) | 12 (60) | 5 (71) |  |
| Composite | 1 (2) | 1 (3) | - | - |  |
| missing | 31 | 17 | 11 | 3 |  |

To characterize the molecular features of HGBCL-NOS, tumors were analyzed using FISH, IHC, gene expression profiling, and sequencing. A summary of all assays performed for this study, as well as data availability for each biopsy, is presented in supplemental Figure 2. FISH analysis identified a *MYC*-rearrangement in 43/92 (47%) biopsies, *BCL2*-rearrangement in 7/86 (8%), and *BCL6*-rearrangement in 11/86 (13%) (Figure 1A and B). The frequency of *MYC*-rearrangement in HGBCL-NOS (47%) was significantly higher than in DLBCL-NOS (6%, *P* < 0.0001) (Figure 1B; supplemental Table 11). Although the frequency of *BCL2-* and *BCL6*-rearrangement was lower in HGBCL-NOS compared to DLBCL-NOS, this is skewed by the higher observed frequency of *MYC*-rearrangement in HGBCL-NOS tumors. By definition, *MYC*-rearranged HGBCL-NOS and DLBCL-NOS cannot harbor a *BCL2-* or *BCL6*-rearrangement otherwise they would be classified as HGBCL-DH. Among *MYC*-rearrangement-negative tumors, there was no difference in the frequency of *BCL2* (16% vs 23%; *P* = 0.51) or *BCL6* (22% vs 30%; *P* = 0.43) rearrangement between HGBCL-NOS and DLBCL-NOS (supplemental Figure 3).

Routine IHC stains were available for the majority of biopsies included in the study (Table 1). MYC protein expression was observed in 34/54 (63%) biopsies, and 59/86 (57%) were positive for BCL2 protein expression. Only 1/49 biopsies with available data were positive for TDT expression. EBV positivity was observed in 1/49 biopsies with available EBER ISH. The median Ki67 proliferation index was 90%. By the Hans algorithm, 68/83 (82%) biopsies had a GCB immunophenotype. By digital gene expression profiling, the distribution of COO subtypes was 59% GCB, 25% ABC, and 16% unclassified. Using the refined COO algorithm, 26/41 (63%) GCB and 5/11 (45%) Unclassified biopsies were reassigned to the DZsig+ subgroup (Figure 1C), totaling 45% of all biopsies, significantly higher than the frequency of DZsig+ tumors observed in DLBCL-NOS (7%; *P* < 0.0001). There was no significant difference in the above molecular characteristics between intermediate and blastoid tumors (Table 1; supplemental Figure 4A).

### Genomic landscape of HGBCL-NOS: structural variants, copy number alterations, and mutations

Using WGS or capture sequencing data available for 66 biopsies, a “cryptic” *MYC*-rearrangement was detected in 4/35 (11%) tumors that were negative for *MYC* break-apart FISH, and a *BCL2*-rearrangement was detected in 3/57 (5%) *BCL2* FISH-negative biopsies (supplemental Figure 5). Accounting for these “cryptic” rearrangements would render a diagnosis of HGBCL-DH-*BCL2* in 6% (4/66) of the sequenced tumors. These biopsies were not excluded from the study as identifying rearrangements that are cryptic to FISH is not currently feasible in routine clinical practice. In addition to the four cryptic *MYC*-rearrangements, a *MYC*-rearrangement partner was identified by sequencing in 87% (23/27) of tumors that were *MYC* FISH-positive. Among identified *MYC*-rearrangements, 80% (25/31) involved an IG locus (22 IGH, 2 IGL, 1 IGK) (Figure 2A). Half of *IGH::MYC* rearrangements had a breakpoint in IGHV or Eμ region, while the remaining half were in a downstream switch acceptor region (Figure 2B). The breakpoint architecture observed in HGBCL-NOS is similar to that of BL and sole *MYC*-rearranged DLBCL-NOS. In contrast, *MYC*-rearrangements in HGBCL-DH-*BCL2* more frequently involve non-IG partners, and for those with *IGH::MYC* rearrangements, the breakpoint is more commonly in a downstream switch acceptor region.^20,33^ Consistently, for tumors with available RNAseq data and where a full-length IGH transcript could be assembled, 84% were predicted to express IGHM (supplemental Figure 6). IGHM expression is also more common among BL and DLBCL-NOS, while expression of a switched IGH isotype is more frequently observed in HGBCL-DH-*BCL2*.^33^ Interestingly, the most frequently used IGH variable gene was IGHV4-34 (20%) (supplemental Figure 6), which is more commonly observed in ABC-DLBCL.^41,42^

**Figure 2.**
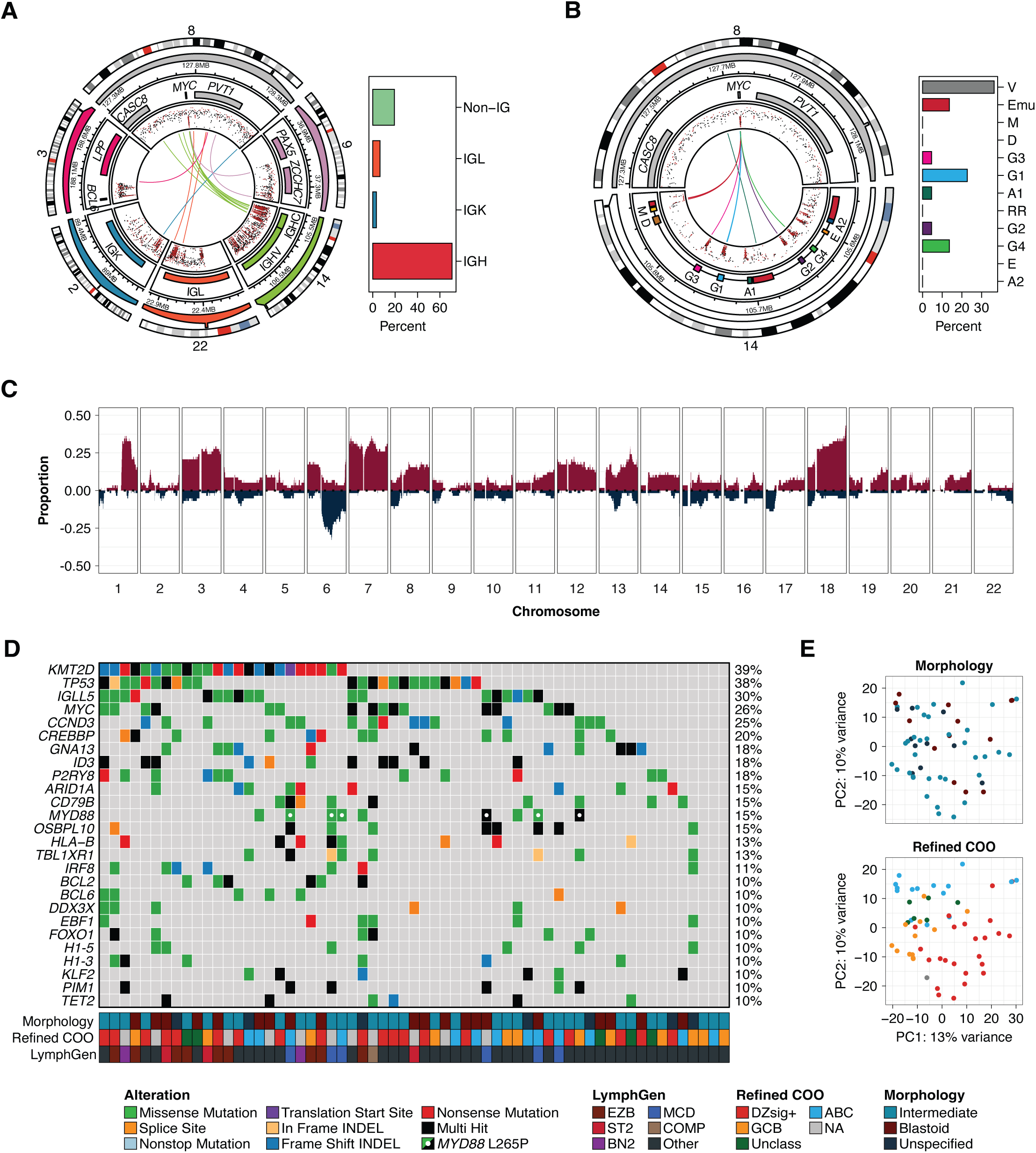
The genetic landscape of HGBCL-NOS. **(A)** The architecture of *MYC*-rearrangements and the observed frequency of IGH, IGK, IGL, and non-IG rearrangement partners. The outermost ring shows the chromosome ideogram followed by a genomic coordinate scale, and a gene coordinate track. Innermost lines show the linkages between *MYC* and the corresponding rearrangement partner for each sample. **(B)** The architecture of *IGH::MYC* rearrangements and the frequency of breakpoints occurring at the indicated regions of the IGH locus. Eμ and 3’ regulatory region (RR) enhancers are included in the gene coordinate track (red). **(C)** Copy number profile of HGBCL-NOS. The proportion of tumors with a copy number gain (red) or deletion (blue) is plotted across each chromosome. **(D)** Oncoplot showing the mutation status of the most frequently mutated genes in HGBCL-NOS for each patient tumor. Genes included in the plot were mutated in at least 10% of HGBCL-NOS samples and identified as a significantly mutated gene HGBCL-NOS, DLBCL-NOS, or BL. **(E)** Principal component analysis of RNA sequencing data.

CNAs were identified from WGS or low pass WGS available for 58 biopsies. Analysis of CNAs using GISTIC2 identified 26 recurrent focal or arm-level CNAs (supplemental Table 12). Included among focal CNAs were amplifications of 2p16.1 (*REL*) and 13q31.3 (*MIR17HG*), and deletions of 1p36.32 (*TNFRSF14*), 6q21 (*PRDM1*), and 13q14.2 (*RB1*) (Figure 2C). The most frequent arm-level CNA was amplification of 18q, which includes *BCL2*. Notably, dysregulation of BCL2 expression by amplification is also commonly observed in ABC-DLBCL.^43^ Blastoid and intermediate morphology tumors displayed similar copy number profiles (supplemental Figure 4C; supplemental Table 13).

Mutations were identified from WGS or WES sequencing available for 61 biopsies. In total, 24 genes were found to be significantly mutated among HGBCL-NOS biopsies. All significantly mutated genes that were present in at least 10% of samples have been previously reported to be recurrently mutated in DLBCL-NOS and/or BL^20^ (supplemental Table 14). The most frequently mutated genes across HGBCL-NOS tumors were *KMT2D* (39%), *TP53* (38%), IGLL5 (30%), *MYC* (26%), *CCND3* (25%), and *CREBBP* (20%), with all other genes mutated in less than 20% of biopsies (Figure 2D). There was no significant difference in the mutational frequency of any recurrently mutated gene between tumors with blastoid or intermediate morphology (supplemental Figure 4B; supplemental Table 15). Consistently, there were limited differentially expressed genes between blastoid and intermediate tumors (supplemental Figure 4D; supplemental Table 16), and by principal component analysis (PCA), tumors did not cluster by morphological subtype (Figure 2E).

To determine if HGBCL-NOS shares similarities with related aggressive B-cell lymphoma entities, comparisons of gene mutational frequencies were made to DLBCL-NOS (n = 341) and BL (n = 63) (supplemental Table 17). Interestingly, *KMT2D* was the most frequently mutated gene among both HGBCL-NOS (39%) and DLBCL-NOS (30%) tumors, observed at similar frequencies in both groups, while being significantly less common in BL (11%; *Q* < 0.01) (Figure 3B). Although comparisons between BL and HGBCL-NOS were limited by sample size, seven genes were more frequently mutated in HGBCL-NOS compared to BL (Q ≤ 0.1), with these mutations almost entirely absent in BL but occurring at similar frequencies in DLBCL-NOS and HGBCL-NOS (Figure 3B). Despite more power in comparisons with the DLBCL-NOS cohort, only three genes were mutated at significantly higher frequency in DLBCL-NOS than in HGBCL-NOS (Figure 3C). In comparison, 24 genes were significantly more frequently mutated in DLBCL-NOS than in BL (supplemental Figure 7). These findings indicate that HGBCL-NOS has a complex genomic landscape overlapping with DLBCL-NOS, suggesting that its heterogeneity, analogous to that of DLBCL-NOS, may be resolved by identifying molecular substructure.

**Figure 3.**
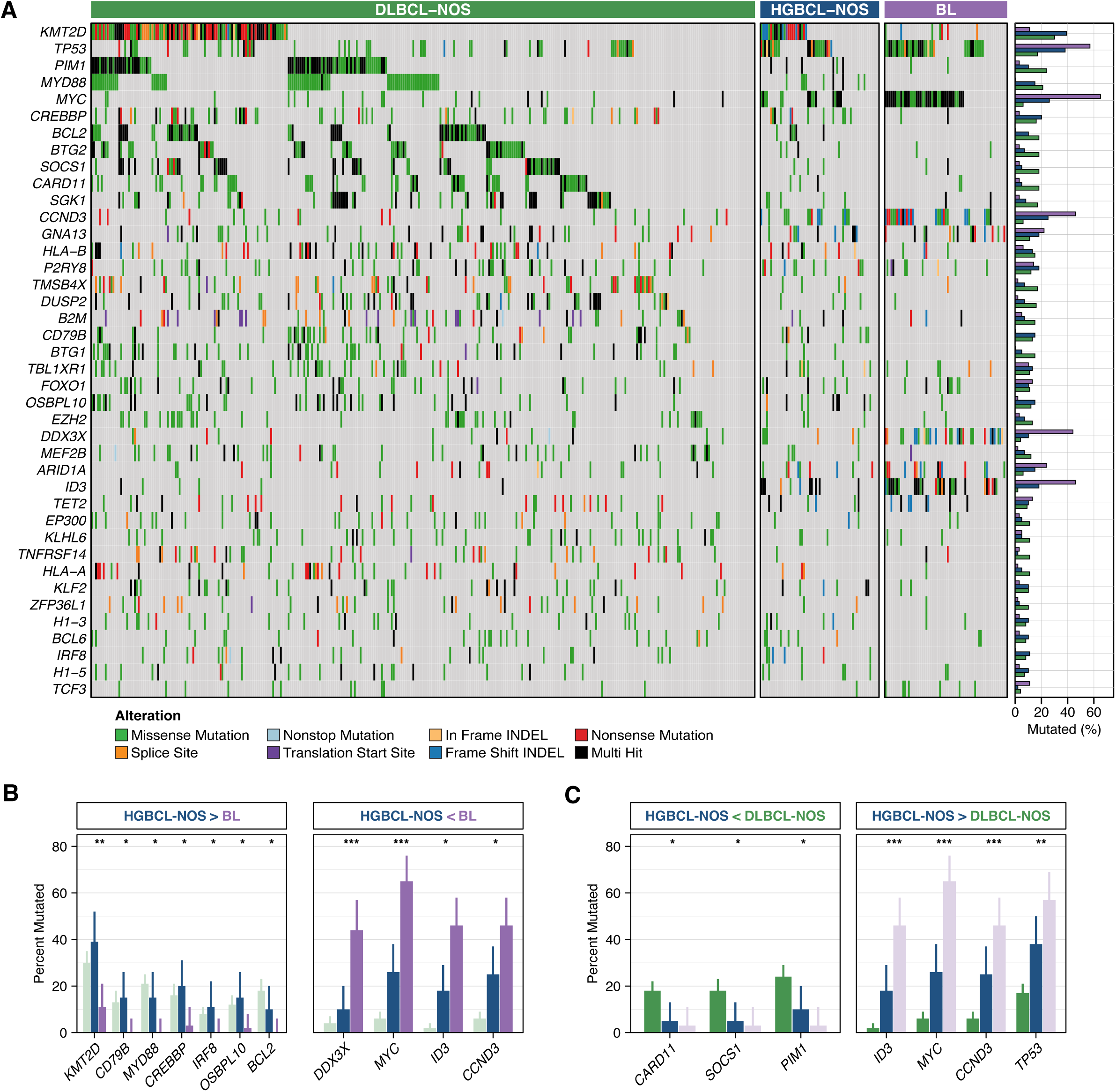
Comparison of the mutational landscape of HGBCL-NOS to DLBCL-NOS and EBV-negative BL. **(A)** Oncoplot showing the mutation status of recurrently mutated genes for 61 HGBCL-NOS, 341 DLBCL-NOS, and 63 BL tumors. HGBCL-NOS and BL tumors were sequenced by WGS or WES, and DLBCL-NOS tumors were sequenced using a targeted capture panel covering 125 genes. Genes included in the plot represent the intersection between 1) genes covered by the targeted capture panel and 2) genes that were identified as significantly mutated in HGBCL-NOS, DLBCL-NOS, or BL and mutated at a frequency of ≥ 10% in at least one of these groups. Nine genes that fulfilled the second criterion were not covered by the capture panel (*EBF1*, *FBXO11*, *IGLL5*, *KIAA1671*, *PCBP1*, *PHF6*, *RFX7*, *SMARCA4*, *TFAP4*). Genes are ordered by their mutational frequency across all groups, and gene mutation frequencies for each group are shown in the bar plots on the right. **(B)** Comparison between HGBCL-NOS and BL, with DLBCL shown for reference. Genes with significantly different mutation frequencies between HGBCL-NOS and BL are displayed. **(D)** Comparison between HGBCL-NOS and DLBCL, with BL shown for reference. Genes with significantly different mutation frequencies between HGBCL-NOS and DLBCL are displayed. P-values were calculated using Fisher’s exact test and adjusted for multiple comparisons using the Benjamini-Hochberg method.

### Molecular classification of HGBCL-NOS

Genetic classification has been an approach to resolve the heterogeneity of DLBCL-NOS.^15,16,18^ Given the similarities between HGBCL-NOS and DLBCL-NOS, we sought to determine if the DLBCL-derived LymphGen genetic classification system^17^ could also identify molecular substructure within HGBCL-NOS. Interestingly, all LymphGen subtypes other than the rare N1 group were represented among HGBCL-NOS tumors (Figure 1B), with the most frequent subtypes being EZB (15%) and MCD (10%). Consistent with MCD being the second most common LymphGen subgroup, mutations in *MYD88* and *CD79B* were each observed in 15% of biopsies, with 6/9 *MYD88* mutated tumors harboring an L265P hotspot mutation (Figure 2D). Although a subset of HGBCL-NOS tumors could be assigned to a LymphGen subgroup, the majority of tumors (66%) remained unclassified (Other) – a significantly higher proportion than in DLBCL-NOS (34%; *P* < 0.00001) (Figure 1B). Notably, 89% of BL could also not be assigned to a LymphGen subgroup (supplemental Figure 8), consistent with BL having molecular features that are distinct from DLBCL-NOS. Of interest, the four most frequently mutated genes in BL (*MYC*, *TP53, CCND3*, *ID3*) also represented mutational features that distinguished HGBCL-NOS from DLBCL-NOS, with all four genes significantly more frequently mutated in HGBCL-NOS than DLBCL-NOS (Figure 3C). However, their frequency did not reach the level seen in BL, with all but *TP53* significantly less frequently mutated in HGBCL-NOS than in BL (Figure 3B). These findings are consistent with HGBCL-NOS comprising multiple tumor groups, with a subset aligning to defined DLBCL-NOS genetic subtypes, and a further subset sharing features with BL. The latter subset may either resemble BL at a genetic level or represent an intermediate group—similar to HGBCL-DH-BCL2—where BL-associated mutations are common among an EZB-DLBCL background.^17,44^

Although originally described in HGBCL-DH-*BCL2*, DZsig expression extends to BL and a subset of GCB DLBCL-NOS.^40^ BLs are universally DZsig+ and have a GCB COO, with other defining features including *MYC*-rearrangement and a lack of high BCL2 expression. In contrast, only ∼15% of GCB DLBCL-NOS are DZsig+, and two-thirds harbor a *BCL2*-rearrangement. Exploring gene expression of HGBCL-NOS using PCA, the top two PCAs clustered samples by refined COO subgroups (Figure 2E), highlighting the utility of refined COO as a framework for organizing the observed genomic heterogeneity. As such, given the high frequency of DZsig expression and *MYC*-rearrangement in HGBCL-NOS, we leveraged the refined COO framework to explore if any HGBCL-NOS tumors bear molecular resemblance to BL.

Among GCB tumors that harbored a *MYC*-rearrangement, DZsig expression was observed in 19/20 (95%) HGBCL-NOS, compared to 12/19 (63%) DLBCL-NOS (*P* = 0.02) (Figure 4A and B). In total, 19/26 (73%) of DZsig+ GCB HGBCL-NOS tumors harbored a *MYC*-rearrangement, compared to only 12/48 (25%) of GCB DZsig+ DLBCL-NOS (*P* = 0.0001) (Figure 4B). Using a previously described random forest classifier, GCB HGBCL-NOS tumors were assigned into BL-like or DLBCL-like genetic subgroups based on their mutational patterns.^20^ Among the 15 DZsig+ GCB tumors with available sequencing, 53% were assigned to the BL-like subgroup, compared to 1/12 (8%) DZsig-GCB tumors (*P* = 0.02; Figure 4C). Within the group of DZsig+ tumors classified as BL-like, 6/8 harbored an *IG::MYC* rearrangement, and all but one tumor had *BCL2* mRNA expression below the cohort median. Similar to BL, all the BL-like tumors were unclassified by LymphGen. In contrast, among the DZsig+ GCB tumors assigned to the DLBCL-like subgroup, 6/7 displayed high levels of *BCL2* mRNA expression, with all six tumors correspondingly positive for BCL2 protein expression by IHC (Figure 4C and D). Five of these tumors harbored *BCL2*-rearrangements, two of which were cryptic to FISH. By LymphGen classification, 5/7 DLBCL-like tumors were classified as EZB, one as ST2, and one as Other (Figure 4D). Of note, only 3/12 (25%) GCB DZsig-tumors were classified into a LymphGen subgroup (supplemental Figure 9).

**Figure 4.**
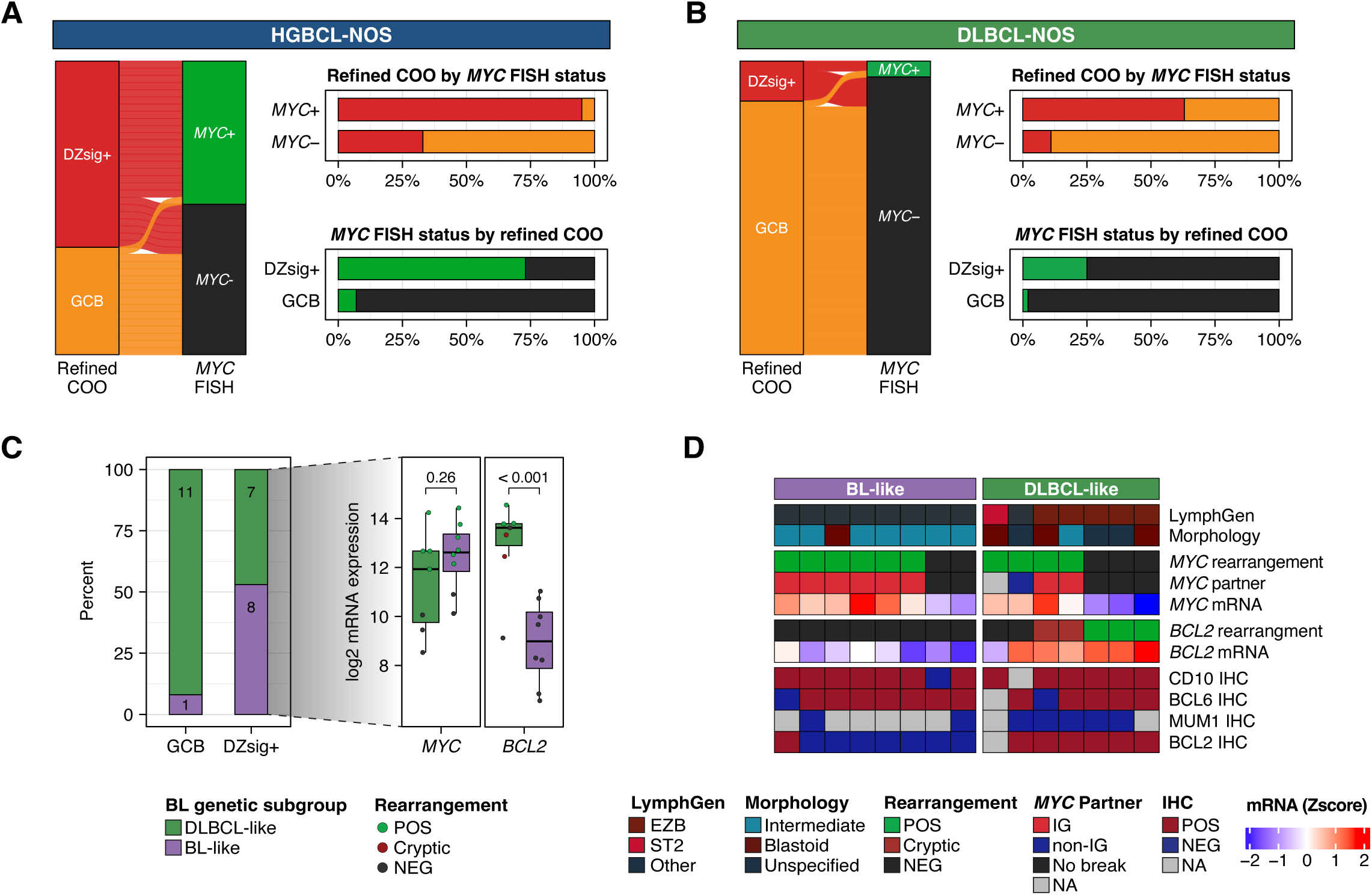
Molecular features of GCB tumors. All analyses are restricted to GCB tumors. For refined COO, the GCB group includes tumors that were negative or indeterminant for DZsig expression. *MYC*- and *BCL2*-rearrangement status was determined by FISH, and *MYC*-rearrangement partners were identified by sequencing. **(A-B)** The relationship between DZsig expression and *MYC*-rearrangement status in **(A)** HGBCL-NOS and **(B)** DLBCL-NOS. Each ribbon in the alluvial plot represents a single biopsy, linking its *MYC*-rearrangement and DZsig expression status (left). Bar plots show the percentage of DZsig+ biopsies stratified by *MYC*-rearrangement status (top right) and the frequency of *MYC*-rearrangement stratified by DZsig expression (bottom right). **(C)** HGBCL-NOS biopsies were assigned into BL-like and DLBCL-like groups using a random forest classifier trained on mutational data to distinguish BL from DLBCL-NOS. The bar plot shows the percentage of tumors classified into DLBCL-like and BL-like groups stratified by DZsig expression status. Box plots display *MYC* and *BCL2* mRNA expression levels in DZsig+ HGBCL-NOS tumors classified as DLBCL-like or BL-like. P values for comparisons between groups were calculated using a t-test. **(D)** Molecular features of BL-like and DLBCL-like DZsig+ HGBCL-NOS tumors. Each column of the heatmap corresponds to a single biopsy. *MYC* and *BCL2* mRNA expression levels are shown as a Z-score relative to all HGBCL-NOS biopsies with available digital gene expression profiling data.

Among ABC HGBCL-NOS tumors, 4/13 (31%) were MCD, while the remainder were unclassified by LymphGen (supplemental Figure 10A). Interestingly, DZsig expression was observed in 5/17 (29%) ABC HGBCL-NOS tumors, which was rarely observed in ABC DLBCL-NOS tumors (1/267; *P* < 0.00001) (supplemental Figure 10B). DZsig expression was only observed among non-MCD ABC tumors, and collectively, 69% of ABC tumors with available digital gene expression and sequencing data were either DZsig+ or MCD (supplemental Figure 10A). Although outcome data was limited in this cohort, the ABC subgroup displayed the worst outcomes, with a 2-year overall survival (OS) of 21% (supplemental Figure 11A). In contrast to DLBCL-NOS^40^, OS was similar in GCB (2y-OS 61%) and DZsig+ (2y-OS 69%) HGBCL-NOS.

However, despite not having DZsig information at the time of treatment, 62% of DZsig+ tumors were treated with an intensified regimen, while all GCB tumors received an R-CHOP-like regimen (supplemental Figure 11B; supplemental Table 18). In aggregate, these results highlight that HGBCL-NOS represents a heterogeneous collection of tumors, and although a subset of tumors have molecular features that align with existing taxonomies, a further subset cannot be confidently classified.

### Centralized Pathology Review

Recognizing the challenges of identifying high-grade morphology, all HGBCL-NOS tumors underwent CPR by the LLMPP panel of hematopathologists. Further details of the CPR process are described in a companion report (Kurz et al). Upon CPR, only 42% of biopsies were confirmed as HGBCL-NOS, and 45% were reclassified as DLBCL-NOS (Figure 5A). The remaining biopsies were reclassified as Burkitt-like lymphoma with 11q aberration (BL11q; n=1), B-cell acute lymphoblastic leukemia/lymphoma (B-ALL; n=1), or BL (n=5), and an additional five biopsies were deemed insufficient for definitive diagnosis. The proportion of tumors confirmed as HGBCL-NOS was similar between those submitted as intermediate (20/51; 39%) and blastoid (16/31; 52%) (*P* = 0.36; Figure 5B). There was no significant difference in clinical characteristics between tumors that were either confirmed as HGBCL-NOS or reclassified as DLBCL-NOS (supplemental Table 19), and no difference in patient overall survival (*P* = 0.94; supplemental Figure 12; supplemental Table 20). There was also no significant difference in molecular characteristics, including the frequency of DZsig expression and *MYC*-rearrangement (Figure 5C). Furthermore, there was no significant difference in the frequency of any recurrently mutated genes (Figure 5D; supplemental Table 21) or CNAs (Figure 5E; supplemental Table 22), as well as no significantly differentially expressed genes that exceeded a log2 fold change of 1 (supplemental Table 23). Consistently, PCA analysis of RNAseq data revealed substantial overlap between CPR diagnostic groups (HGBCL-NOS and DLBCL-NOS) (Figure 5F).

**Figure 5.**
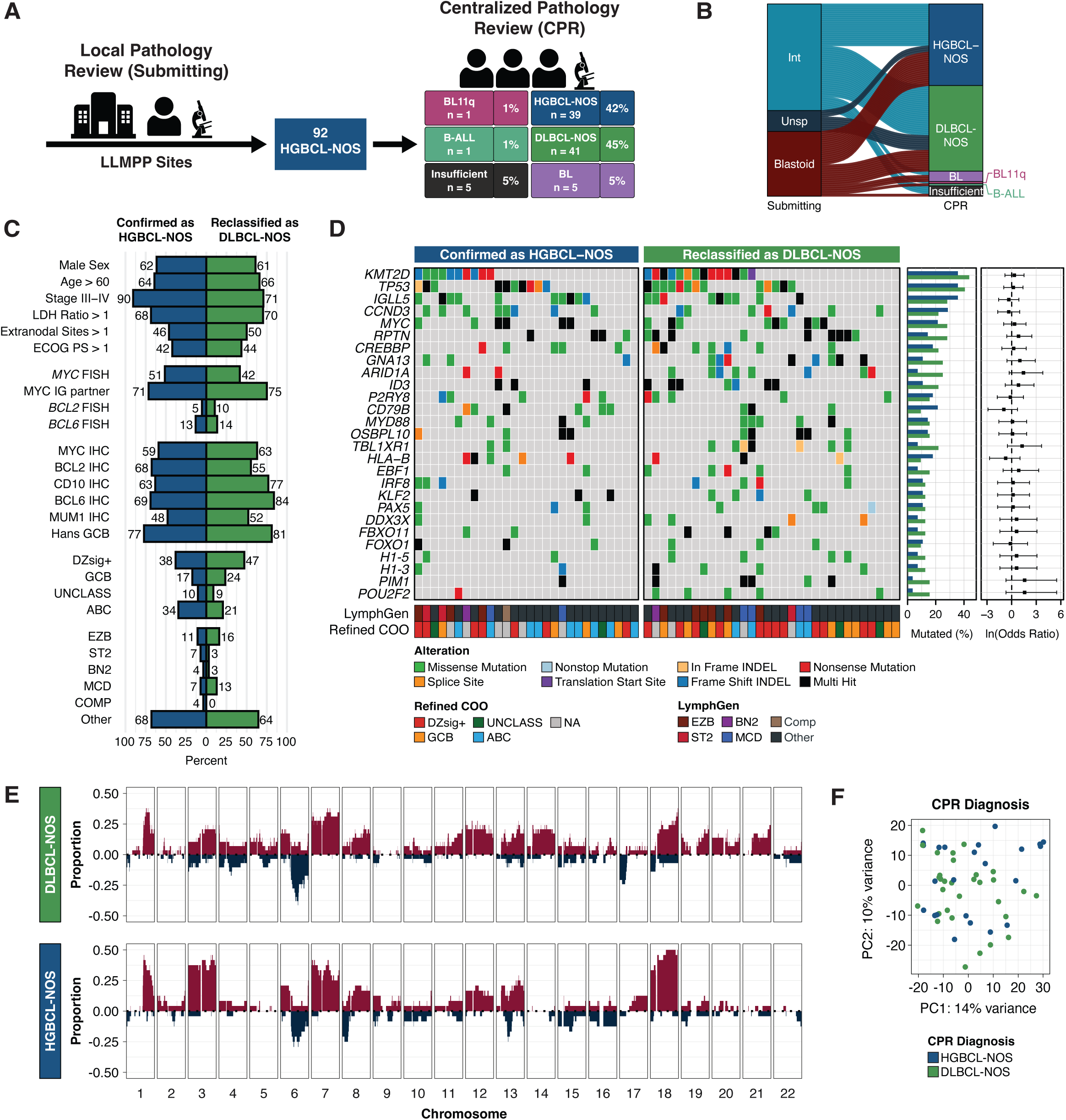
Comparison of tumors by centralized pathology review assignment. **(A)** Biopsies were initially reviewed locally by LLMPP hematopathologists at each site to confirm HGBCL-NOS diagnosis and assign a morphological subtype. All biopsies then underwent centralized pathology review by a panel of LLMPP hematopathologists. Biopsies were either confirmed as HGBCL-NOS, reclassified as another lymphoma entity or deemed insufficient for diagnosis. **(B)** Concordance of submitted morphological subtype and centralized pathology review diagnosis. Each ribbon in the alluvial plot represents a single biopsy, linking its morphological subtype assigned by the submitting site to the centralized review diagnosis. **(C)** Clinical and molecular features in tumors confirmed as HGBCL-NOS or reclassified as DLBCL-NOS. Comparisons of clinical, FISH, and IHC data were performed using Fisher’s exact test, while refined COO and LymphGen classifications were compared by χ² test. No comparisons were statistically significant. **(E)** Comparison between the mutational landscape of tumors confirmed as HGBCL-NOS or reclassified as DLBCL-NOS. Genes included in the oncoplot were mutated in at least 10% of HGBCL-NOS samples and identified as a significantly mutated genes in HGBCL-NOS, DLBCL-NOS, or BL. Gene mutation frequencies for each group are shown in the bar plots on the right, with odds ratios and 95% confidence intervals for each gene comparison shown in the forest plots. **(F)** Copy number profiles of tumors confirmed as HGBCL-NOS or reclassified as DLBCL-NOS. The proportion of tumors with a copy number gain (red) or deletion (blue) is plotted across each chromosome. **(F)** Principal component analysis of RNA sequencing data.

## Discussion

Our characterization of a sizeable cohort of HGBCL-NOS tumors gathered across the LLMPP consortium, incorporating centralized pathology review and comprehensive molecular profiling, aimed to define common and distinct biological features of this disease. Although the frequency of *MYC*-rearrangement and DZsig-positivity, both observed in approximately half of tumors, was distinct from both BL and DLBCL-NOS, the study revealed HGCBL-NOS to be a highly heterogeneous disease. Centralized review highlighted the challenge of using morphology as a defining feature, with over half of HGBCL-NOS tumors reassigned to other categories, predominantly DLBCL-NOS. However, this reclassification did not reduce the molecular heterogeneity observed within HGBCL-NOS, with no differences in molecular features between confirmed and reclassified tumors.

HGBCL-DH-*BCL2* encompasses tumors with a spectrum of morphologies.^2,3^ In an analogous fashion, given the rarity of HGBCL-NOS and the absence of a unified set of molecular features, one way of considering this category is as comprising tumors that represent morphological extremes of established categories. At the gene expression level, the refined COO groupings encapsulated the majority of variation seen across HGBCL-NOS, marking this system as a useful method for parsing tumors. Using gene expression profiling to assign refined COO subtypes, 45% were DZsig+, 22% GCB, and 25% ABC, with 9% being unclassified. Within the DZsig+ group, most of the tumors had molecular features strongly aligned with either BL or HGBCL-DH-*BCL2*, a minority of which harbored cryptic rearrangements that would have led to definitive membership. These results are consistent with those seen in the pediatric *MYC*-rearranged lymphoma, where most were DZsig+ and had mutational profiles consistent with BL, irrespective of high-grade or DLBCL morphology.^45^ Further, approximately one-third of ABC HGBCL-NOS tumors were assigned to the MCD genetic subtype. However, many tumors cannot be resolved using this approach, representing likely rare biology not observed in established categories. One such example is the high proportion of ABC HGBCL-NOS expressing DZsig, a phenotype rarely seen in DLBCL-NOS, which requires further exploration.

Our study was not designed or powered to determine the outcomes of HGBCL-NOS. The absence of a standard treatment and the inclusion of tumors from across the LLMPP consortium led to variability in treatment regimens. Nonetheless, consistent with Zayac et al^9^, the poorest outcomes were observed among ABC tumors. Although treatment escalation, as used in BL and HGBCL-DH-*BCL2*, may be appropriate for DZsig+ HGBCL-NOS, agents with efficacy in ABC DLBCL-NOS, such as polatuzumab vedotin and BTK inhibitors,^46,47^ may be the route to improving outcomes in ABC HGBCL-NOS. The challenges associated with morphological classification and the rarity of these tumors supports the inclusion of these patients in DLBCL-NOS trials, particularly those incorporating biomarker-guided treatment arms and/or retrospective molecular characterization.^9^

In conclusion, HGBCL-NOS represents a heterogeneous collection of aggressive B-cell lymphomas. Beyond detecting recurrent rearrangements to exclude HGBCL-DH-*BCL2*, more extensive molecular profiling of these high-grade morphology tumors is encouraged as the majority can be aligned to established pathology categories and/or genetic groups using this information. Finally, further work is needed to understand the origins of the morphological variance along with the biology of the remaining tumors that cannot currently be molecularly realigned.

## Data Availability

The results published here are in part based upon data generated by the Cancer Genome Characterization Initiative (phs000235), Burkitt Lymphoma Genome Sequencing Project (phs000527) and Non-Hodgkin Lymphoma - Diffuse Large B-Cell Lymphoma (NHL-DLBCL; phs000532), developed by the NCI. The data used for this analysis are available at https://www.ncbi.nlm.nih.gov/projects/gap/cgi-bin/study.cgi?study_id=phs000235.v21.p6. All additional sequencing data generated for this study will be deposited in the European Genome-phenome Archive (EGA).

## Acknowledgements

This study was supported by grant 1P01CA229100 from the National Cancer Institute, the Canadian Cancer Society Research Institute (704848 and 705288), and Terry Fox Research Institute (TFRI) Program Project Grants (1061, 1108). It was also supported by a Large Scale Applied Research Project funded by Genome Canada (13124), Genome BC (271LYM), Canadian Institutes of Health Research (CIHR) (GP1-155873), the British Columbia Cancer Foundation (BCCF) and the Provincial Health Services Authority (PHSA). BC is supported by a CIHR Canada Graduate Scholarship Doctoral Award. RDM and CS are supported by Michael Smith Foundation for Health Research Career Investigator Awards. DWS is supported by a Michael Smith Foundation for Health Research, Health Professional Investigator Award.

The results published here are in part based upon data generated by the Cancer Genome Characterization Initiative (phs000235), Burkitt Lymphoma Genome Sequencing Project (phs000527) and Non-Hodgkin Lymphoma – Diffuse Large B-Cell Lymphoma (NHL-DLBCL; phs000532), developed by the NCI. The data used for this analysis are available at https://www.ncbi.nlm.nih.gov/projects/gap/cgi-bin/study.cgi?study_id=phs000235.v21.p6.

## Author contributions

Designed research: B.C., J.R.C., G.O., R.D.M., L.M.R and D.W.S.

Performed research: B.C., S.B-N., W.A., G.W.S., P.F., M.B., B.M., J.R.C., G.O., A.R., E.C., C.A., T.C.G., P.W.R., J.Y.S., G.I., S.L.O., E.S.J., D.D.W., W.C.C., H.H., K.B., K.F., J.D., S.P., G.W., A.L.M., A.J.M., A.L.F., L.R.M., D.W.S.

Provided materials and data: W.A., G.W.S., P.F., J.R.C., G.O., A.R., E.C., C.A., T.C.G., P.W.R., J.Y.S., G.I., S.L.O., J.I., K.J.S., C.S., L.M.R., D.W.S.

Analyzed data: B.C., L.K.H., J.W., W.A., L.M.S., R.D.M., L.M.R., D.W.S.

Wrote the manuscript: B.C. and D.W.S.

Edited and approved the manuscript: all authors.

