## Supplemental Methods and Figures for "High-grade B-cell lymphoma, not otherwise specified: an LLMPP study"

#### Study cohort

Participating institutions submitted paraffin blocks, immunophenotypic and genetic results, and, when available, clinical records for cases diagnosed as high-grade B-cell lymphoma, not otherwise specified (HGBCL-NOS), according to the WHO revised 4th edition criteria.[1] Bone and bone marrow biopsies, as well as fine needle aspirates, were excluded. Additionally, cases of high-grade B-cell lymphoma with *MYC* and *BCL2* and/or *BCL6* rearrangements (HGBCL-DH) were not included. However, a subset of cases lacked clinical FISH data, preventing definitive exclusion of HGBCL-DH. For cases with sufficient material available for tissue microarray (TMA) inclusion, FISH analysis for *MYC*, *BCL2*, and *BCL6* was performed (described below). Cases confirmed as HGBCL-DH, along with those missing FISH data that could not be included in the TMA, were excluded from the final cohort. Other exclusion criteria included patient age under 18 years, known human immunodeficiency virus (HIV) infection, post-transplant lymphoproliferative disorders, and primary central nervous system lymphomas. In instances where a primary biopsy was unavailable, a relapse biopsy was analyzed for seven patients. Additionally, five patients had a prior diagnosis of indolent lymphoma.

Each biopsy was locally reviewed by an LLMPP hematopathologist to confirm HGBCL-NOS diagnosis and to assign a cytological subtype. Cytological subtypes included: (1) Intermediate between diffuse large B-cell lymphoma (DLBCL) and Burkitt lymphoma (BL) ("intermediate cytology"): defined as a predominance of medium-sized cells with round to oval nuclei, small basophilic nucleoli, but more irregularity in shape and/size than those of classic BL; (2) Blastoid: defined as a predominance of cells with finely dispersed chromatin, sparse cytoplasm, with inconspicuous nucleoli  as may be seen in lymphoblastic lymphoma; (3) Other: used for rare cases not fitting intermediate or blastoid cytology (e.g. cases resembling small immunoblasts). The final cohort consisted of 92 patients with an available HGBCL-NOS biopsy.

### Comparison cohorts

Data from two previously described cohorts were used to compare the molecular features of HGBCL-NOS with Burkitt lymphoma (BL) and diffuse large B-cell lymphoma, not otherwise specified (DLBCL-NOS). The DLBCL-NOS cohort was drawn from a population-based cohort of adult patients (≥ 18 years) diagnosed with a *de novo* tumor of DLBCL morphology between 2005 and 2010 in British Columbia, Canada.[2] Detailed description of this cohort is available in Alduaij et al.[2] In brief, diagnosis was made according to WHO revised 4$^{th}$ edition criteria.[1] Patients with positive HIV serology, post-transplant lymphoproliferative disorder, a prior history of indolent lymphoma, or composite lymphoma were excluded. Data available for this cohort included FISH (*MYC*, *BCL2*, *BCL6*), IHC (MYC, BCL2, CD10, BCL6, MUM1), and DLBCL90 gene expression profiling. All data were generated similarly to as described below for the main study cohort. Analyses were restricted to 781 DLBCL-NOS biopsies with sufficient FISH data to exclude a diagnosis of HGBCL-DH. A summary of the full cohort and the inclusion of each biopsy is provided in supplemental Figure 1A.

The BL cohort was drawn from the Burkitt Lymphoma Genome Sequencing Project (BLGSP).[3] Inclusion was restricted to adult patients (≥ 18 years) with an EBV-negative biopsy to align with the HGBCL-NOS cohort inclusion criteria and avoid EBV status as a confounding factor, as EBV-positive BL is associated with distinct molecular features.[3,4] Inclusion was further restricted to those with available whole genome sequencing (WGS), and in which a *MYC* rearrangement was detected. A total of 63 biopsies met these criteria. A summary of the full BLGSP cohort and the inclusion of each biopsy is provided in supplemental Figure 1B. WGS data for the BL cohort is deposited in dbGaP (Study Accession: phs000527.v18.p6).

### FISH and IHC

Tissue microarrays (TMAs) were constructed from duplicate 0.6 mm cores from formalin-fixed paraffin-embedded (FFPE) blocks. Fluorescent *in situ* hybridization (FISH) was performed on TMA sections using commercially available Metasystems XL break apart probes for *MYC* (D-6030-100-TC), *BCL2* (D-6018-100-OG) and *BCL6* (D-6016-100-OG).

Images were captured using a Metafer CoolCube 1 camera and Metafer software (version 3.11.8). Two independent observers scored 100 nuclei per biopsy. For biopsies with discrepant scores, consensus was reached by a third observer. Rearrangement was defined as a break-apart signal in ≥5% of cells. Immunohistochemistry (IHC) was performed on TMA sections using the Dako Omnis platform for MYC (EP121, Biocare), BCL2 (E17, Abcam; and BCL2/124, Dako), CD10 (DAK-CD10, Dako), BCL6 (PG-B6p, Dako), MUM1 (MUM1p, Dako), CD20 (L26, Dako), TDT (EP266, Dako), and KI67 (MIB-1, Dako). EBER status was determined by EBER ISH (Dako). IHC was independently scored by two expert hematopathologists, and biopsies with discordant scores were further evaluated by a third hematopathologist to reach consensus. For biopsies not included on TMAs, clinical IHC and FISH data provided by the submitting institution were used when available.

**Digital gene expression profiling**

Total nucleic acids (TNA) were extracted from FFPE biopsies using the ALINE FFPE GenePure Purification Kit, automated on the Hamilton Nimbus robot. RNA was isolated by DNAse treatment of TNA. Digital gene expression profiling (GEP) was performed on RNA using the DLBCL90 assay on the nCounter platform (Nanostring), as previously described.[2,5]

**DNA Library construction and sequencing**

The preservation of each tumor sample (FFPE or fresh frozen) is indicated in supplemental Table 10. For FFPE samples, total nucleic acid (TNA) was extracted using the ALINE FFPE GenePure Purification Kit. When ≥300 ng of FFPE extracted TNA was available, TNA was also treated with S1 nuclease to reduce the rate of chimeric fragments derived from single-stranded DNA and overhangs.[6] DNA was isolated from TNA by RNase treatment, and paired-end DNA libraries for sequencing were generated using a PCR-based protocol as previously described.[7] For fresh frozen tissues, DNA was extracted and subjected to library construction as previously described.[8]

HGBCL-NOS biopsies with available fresh frozen tissue underwent whole-genome sequencing (WGS), while FFPE biopsies were analyzed using whole-exome sequencing (WES), targeted capture sequencing, and low-pass WGS, prioritized in this order based on nucleic acid availability. Sequencing data availability for FFPE biopsies varied due to sequencing failures and/or insufficient nucleic acids. Whole exome capture was performed using the SureSelectXT Human All Exon V6 + UTR (Agilent) kit. For targeted capture sequencing, hybridization capture was performed with a panel created by IDT or Twist Biosciences, as previously described.[9] This panel was designed to capture the regions around *MYC* (Chr8:126262754-129664254), *BCL2* (Chr18:63077345-63128767), *BCL6* (Chr3:187721376-188032725), and recurrent rearrangement partner loci: IGH (Chr14:105546276-106879844), IGK (Chr2:88686728-89330586), IGL (Chr22:22329967-23061520), *PAX5* (Chr9:37034478-37465410). Libraries were sequenced on an Illumina HiSeq2500, HiSeqX, or NovaSeq with 150 bp paired end reads.

**Target sequencing of the DLBCL-NOS cohort**

Targeted capture sequencing of 125 genes was performed for 341 DLBCL-NOS biopsies with adequate nucleic acids. The panel was designed to cover genes that are recurrently mutated in aggressive B-cell lymphomas ("LySeqST"; supplemental Table 8). DNA was isolated from FFPE material, and libraries were constructed and sequenced as described above.

**Alignment and variant calling**

WGS, WES, LySeqST targeted sequencing, and low-pass WGS were aligned to the GRCh37 reference genome using bwa-mem (v0.7.17)[10]. Somatic variants were identified with the SLMS-3 pipeline, an ensemble variant calling approach which uses Strelka2[11], LoFreq[12], Mutect2[13], and SAGE. Only variants called by three or more variant callers are passed. Somatic variants were annotated with VCF2MAF and the Ensembl Variant Effect Predictor[14]. The GnomAD reference was used to remove germline variants with a population allele frequency > 1e-4, and further filtered against a custom blacklist of variants that recur when tumors are run without, but not with, a matched normal.

Significantly mutated genes were identified using dNdScv[15], MutSig2CV[16], and OncodriveFML[17]. LymphGen classification[18] was performed on variant calls using somatic mutations and *BCL2/BCL6*-rearrangement data. Variant calls were further used to assign tumors into BL-like and DLBCL-like genetic subtypes using a previously described random forest classifier.[3] Tumors classified to the IC-BL and DGG-BL subgroups were collapsed into a single BL-like group. Copy number alterations (CNAs) were identified from WGS or low-pass WGS using Control-FREEC[19], and recurrent CNAs were identified using GISTIC2[20].

Targeted capture sequencing data for identification of structural variants (SVs) was aligned to the hg38 reference genome using bwa-mem (v0.7.17). SVs involving *MYC* and *BCL2* were identified by an ensemble approach using GRIDSS2[21] and Manta[22], with VCF files output by these tools intersected using the BioConductor package StructuralVariantAnnotation[23]. SVs were filtered out unless they met a minimum variant allele frequency of 0.05 and/or passed manual curation in IGV[24]. WGS data was analyzed using the same pipeline as capture data, and mapped to the hg38 genome build with CrossMap.[25] Circular plots were generated with the R package Circlize.[26] Bioinformatic pipelines are available as part of the LCR Modules repository. Genome coverage was estimated using Picard CollectWgsMetrics. Coverage of exome and targeted sequencing data were estimated with Picard CollectHsMetrics.

**RNAseq analysis**

RNA was isolated by DNase treatment of TNA extracted from FFPE material, as described above. RNAseq libraries were constructed with a ribodepletion method and sequenced with a target of 100M reads per library. RNAseq data were pseudoaligned to the Gencode version 33 transcriptome reference (hg38) with Salmon[27] and normalized with DESeq2[28] and variance stabilizing transformation (VST). To identify functional Ig rearrangements, RNAseq data were analyzed with MiXCR using LCR modules with default settings, mapping predicted rearrangement sequences to the NCBI IgBLAST reference version 1.17.1.

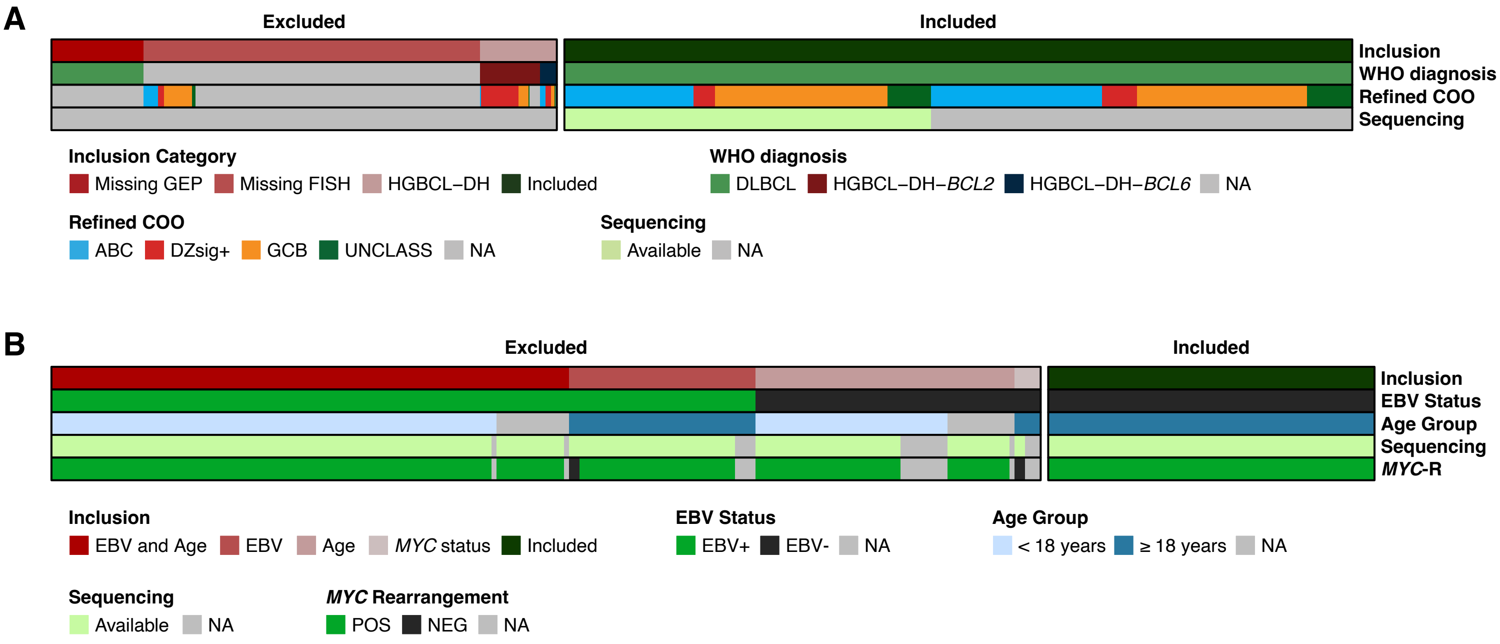


**Supplemental Figure 1. Summary of DLBCL-NOS and BL comparison cohorts. (A)** The DLBCL-NOS comparison cohort consisted of 781 biopsies drawn from a population-based cohort of 1148 adult patients diagnosed with a de novo tumor of DLBCL morphology. HGBCL-DH, as well as biopsies with insufficient FISH to rule out a diagnosis of HGBCL-DH, were excluded. Biopsies were further restricted to those with available digital gene expression profiling data (DLBCL90). **(B)** The BL comparison cohort consisted of 63 biopsies drawn from a cohort of 254 BL biopsies. Inclusion in this study was limited to EBV-negative BL tumors from patients aged 18 or older. Further selection criteria required tumors to have available WGS and a detected *MYC*-rearrangement.

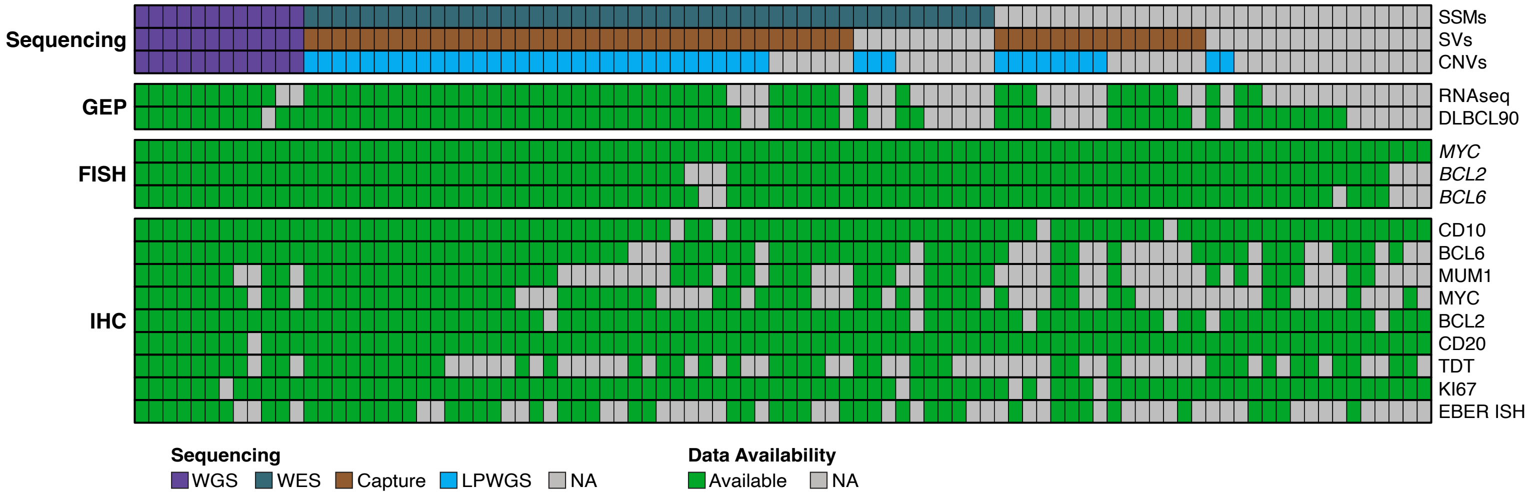


**Supplemental Figure 2. Summary of data availability for the HGBCL-NOS cohort.** Simple somatic mutations (SSMs) were identified from whole genome (WGS) or whole exome sequencing (WES). MYC and BCL2 structural variants (SVs) were identified from WGS or hybridization capture sequencing using a panel designed to capture the regions around *MYC*, *BCL2*, and recurrent rearrangement partner loci. Copy number alterations (CNAs) were detected from WGS or low pass WGS. Digital gene expression profiling using the DLBCL90 assay was used to determine COO and refined COO subtypes. All other transcriptome analyses were performed using RNA sequencing data. *MYC*, *BCL2*, and *BCL6* rearrangements were detected by FISH. EBV status was determined by EBER ISH. Proliferation index scores were determined by KI67 IHC. IHC stains were also performed for CD10, BCL6, MUM1, MYC, BCL2, CD20, and TDT.

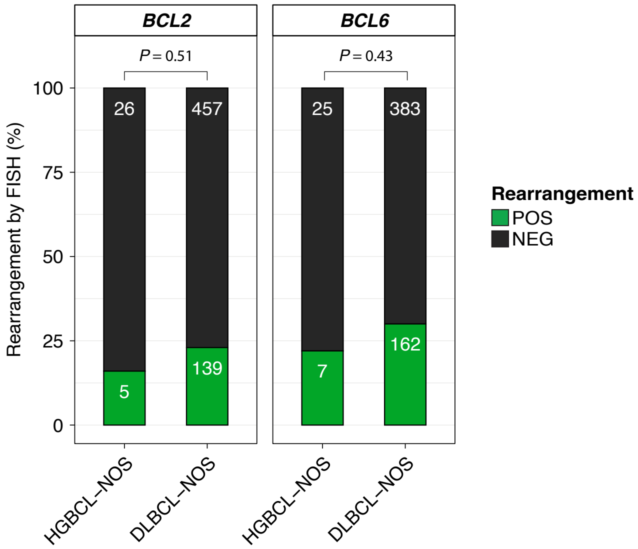


**Supplemental Figure 3. Comparison of *BCL2*- and *BCL6*-rearrangement frequencies between HGBCL-NOS and DLBCL-NOS biopsies negative for *MYC*-rearrangement.** Rearrangement status was determined by FISH. P values were calculated using Fisher's exact test.

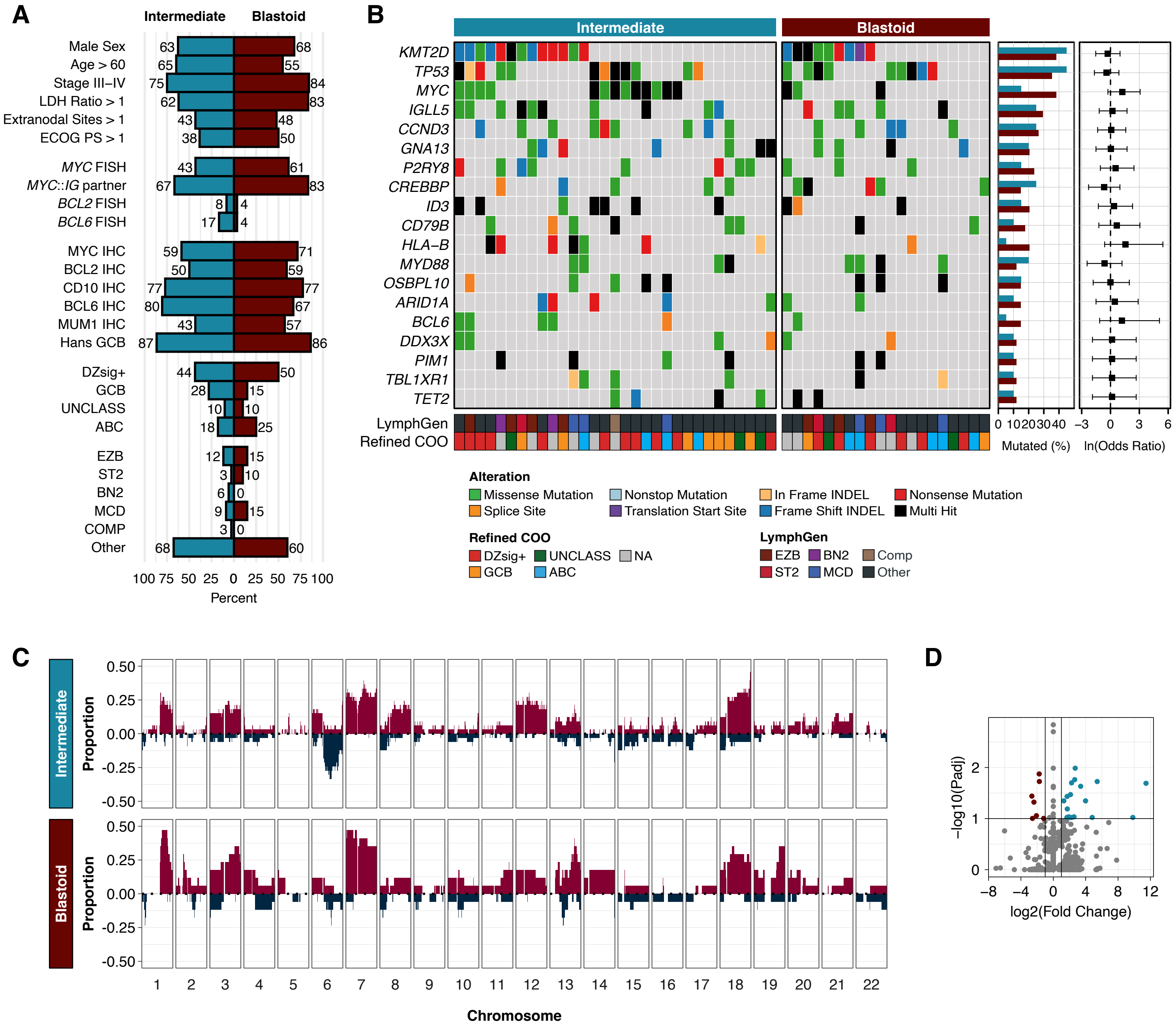


**Supplemental Figure 4. Comparison of the genetic landscape of HGBCL-NOS tumors with intermediate or blastoid morphology. (A)** Clinical and molecular features in HGBCL-NOS tumors with intermediate or blastoid morphology. Comparisons of clinical, FISH, and IHC data were performed using Fisher's exact tests, while refined COO and LymphGen classifications were compared by $\chi^2$ tests. No comparisons were statistically significant. **(B)** Comparison between the mutational landscape of HGBCL-NOS tumors with intermediate or blastoid morphology. Genes included in the oncoplot were mutated in at least 10% of HGBCL-NOS samples and identified as a significantly mutated gene in HGBCL-NOS, DLBCL-NOS, or BL. Gene mutation frequencies for each group are shown in the bar plots on the right, with odds ratios and 95% confidence intervals for each gene comparison shown in the forest plots. **(C)** Copy number profiles of HGBCL-NOS tumors with intermediate or blastoid morphology. The proportion of tumors with a copy number gain (red) or deletion (blue) is plotted across each chromosome. **(D)** Principal component analysis of RNA sequencing data.

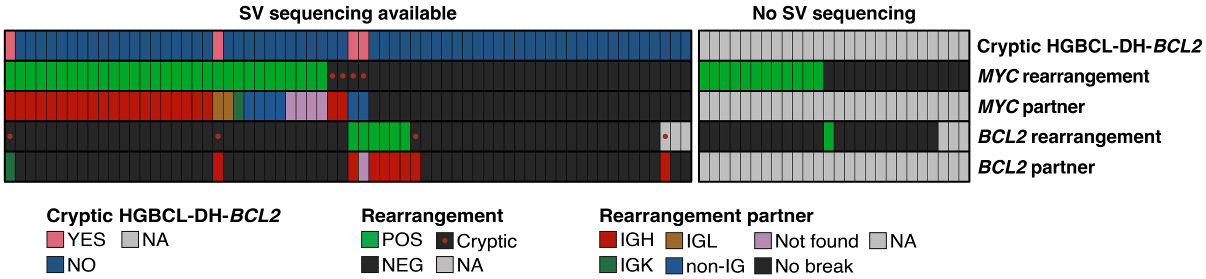


**Supplemental Figure 5. Summary of MYC and BCL2 SV sequencing.** The heatmap is divided into groups based on the availability of sequencing data. Rearrangement status, as determined by FISH, is shown for *MYC* and *BCL2*. Rearrangement partners were determined by sequencing. Tumors with negative or missing *MYC* or *BCL2* FISH results in which a rearrangement was identified by sequencing are marked with a red dot.

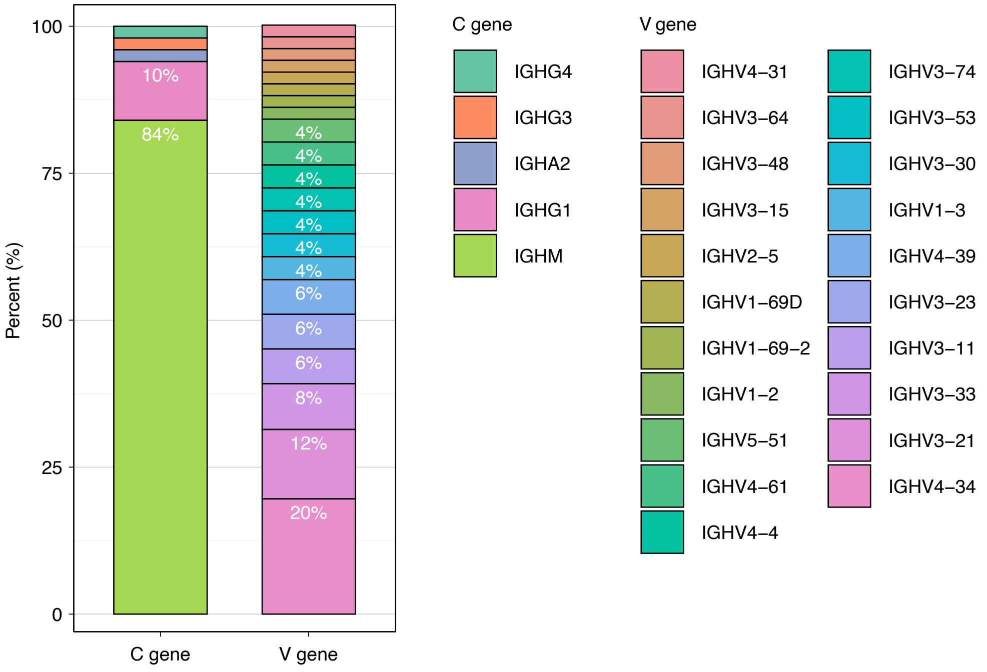


**Supplemental Figure 6. Immunoglobulin heavy chain constant and variable gene usage in HGBCL-NOS.** Full length immunoglobulin transcripts of the dominant clone from each tumor were predicted from RNAsequencing data using MiXCR.

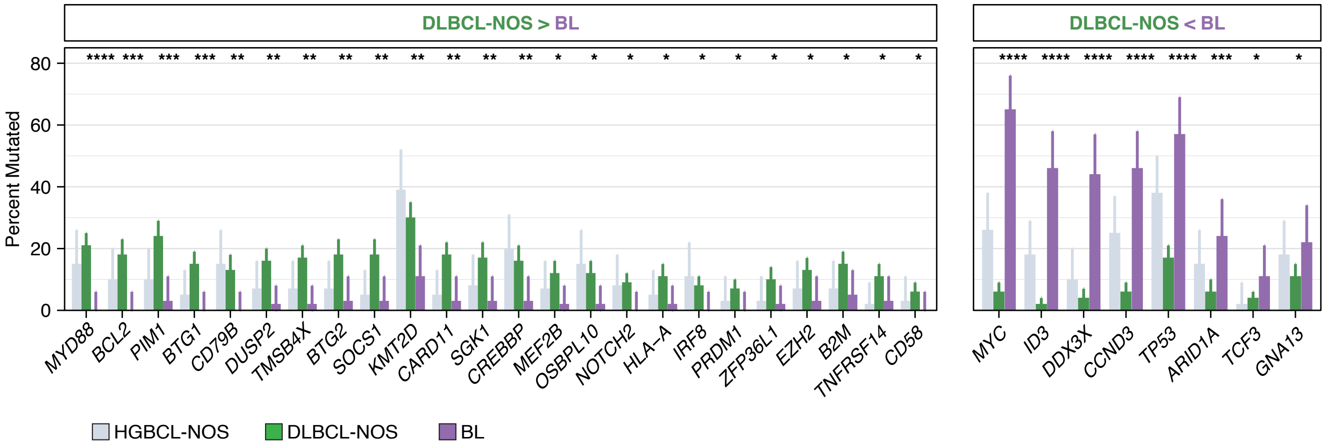


**Supplemental Figure 7. Comparison between the mutational landscapes of DLBCL-NOS and BL.** Bar plots highlighting genes that were significantly differentially mutated between DLBCL-NOS (n = 341) and BL (n = 63). Genes that were more frequently mutated in DLBCL-NOS are shown in the left panel, and genes more mutated in BL are show in the right panel. BL tumors were sequenced by WGS, and DLBCL-NOS tumors were sequenced using a targeted capture panel covering 125 genes. Genes included for comparison represented the intersection between 1) genes covered by the targeted capture panel and 2) genes that were identified as significantly mutated in HGBCL-NOS, DLBCL-NOS, or BL and mutated at a frequency of ≥10% in at least one of these groups. Nine genes that fulfilled the second criteria were not covered by the capture panel (*EBF1*, *FBXO11*, *IGLL5*, *KIAA1671*, *PCBP1*, *PHF6*, *RFX7*, *SMARCA4*, *TFAP4*). P values were calculated using Fisher's exact test and adjusted for multiple comparisons using the Benjamini-Hochberg method.

**Q* < 0.1; ***Q* < 0.01; ****Q* < 0.001; *****Q* < 0.0001.

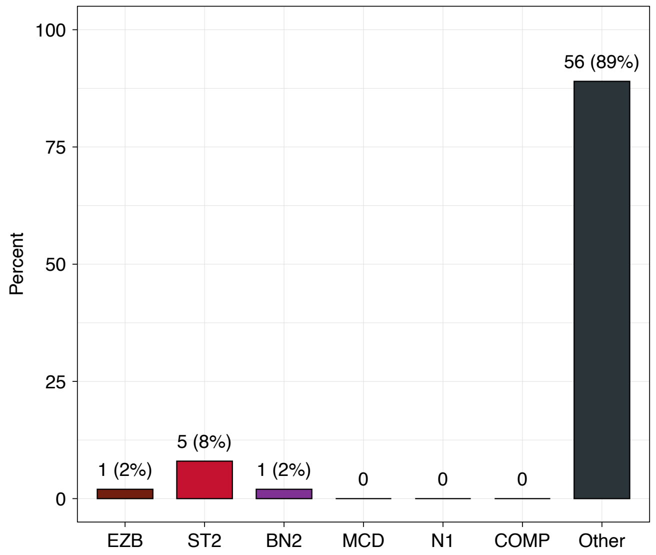


**Supplemental Figure 8. LymphGen classification in BL.** LymphGen classification was performed using somatic mutations and *BCL2/BCL6*-rearrangement data. Somatic mutations and rearrangements were identified by WGS.

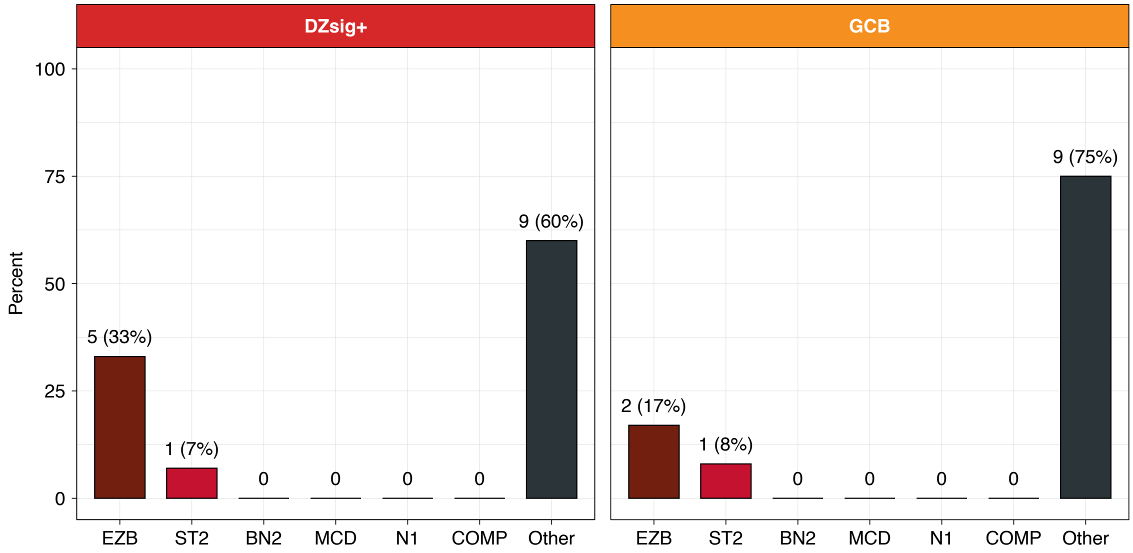


**Supplemental Figure 9. LymphGen classification in GCB HGBCL-NOS.** LymphGen classification was performed using somatic mutations and *BCL2/BCL6* rearrangement data. Somatic mutations were identified in WGS or WES data and rearrangements were identified by FISH. Only GCB tumors are included in the DZsig+ group. The GCB group refers to GCB tumors that were negative or indeterminant for DZsig expression.

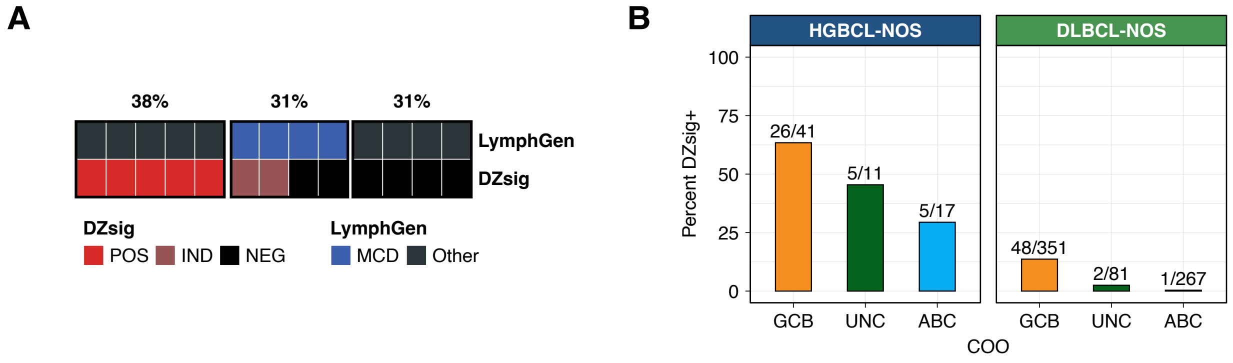


**Supplemental Figure 10. Molecular features of ABC HGBCL-NOS. (A)** LymphGen subtype and DZsig expression status for ABC HGBCL-NOS tumor with available sequencing data. **(B)** The percentage of HGBCL-NOS (left) and DLBCL-NOS (right) tumors that are positive for DZsig expression within each COO subgroup.

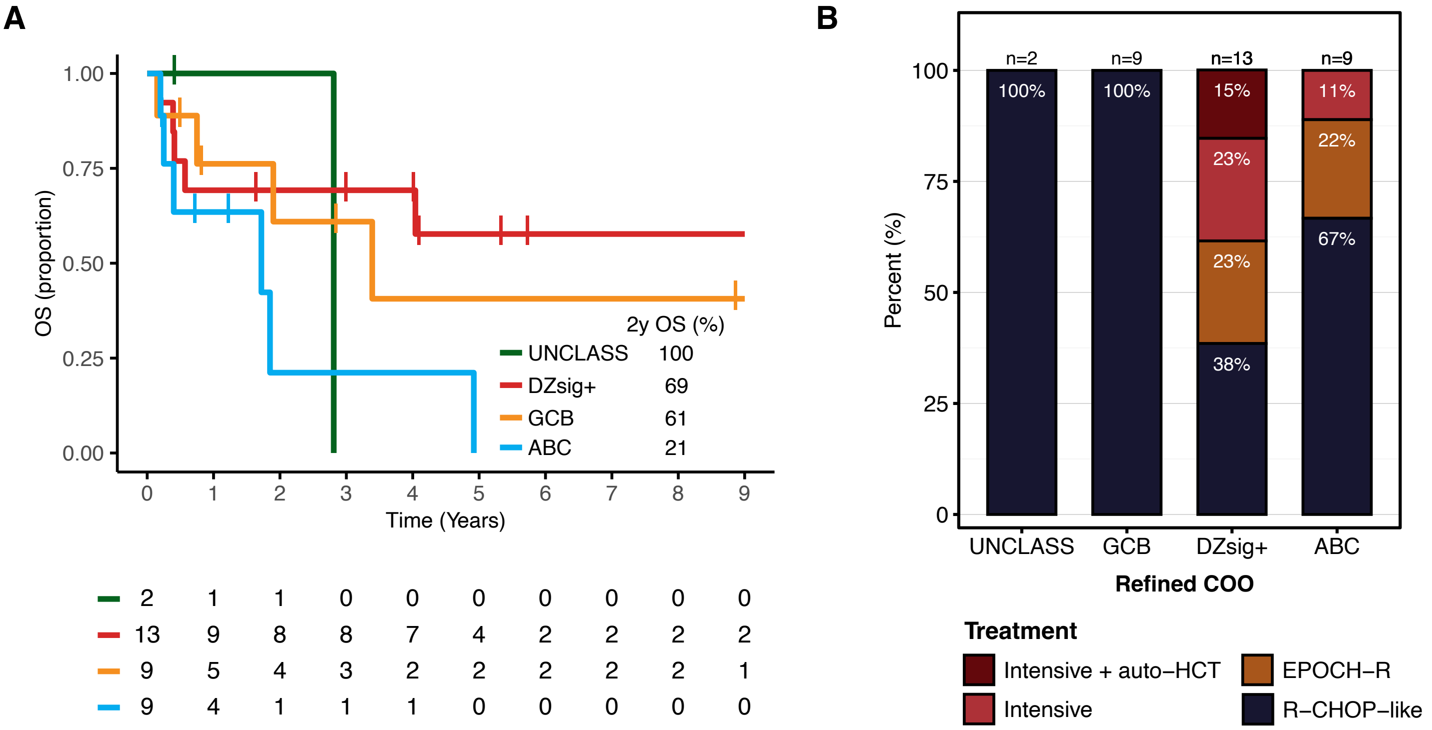


**Supplemental Figure 11. Outcomes in HGBCL-NOS by refined COO subgroups. (A)** Overall survival (OS) by refined COO subgroups. OS was determined from the date of diagnosis to the date of last follow-up or death from any cause. **(B)** Treatment regimen stratified by refined COO subgroups.

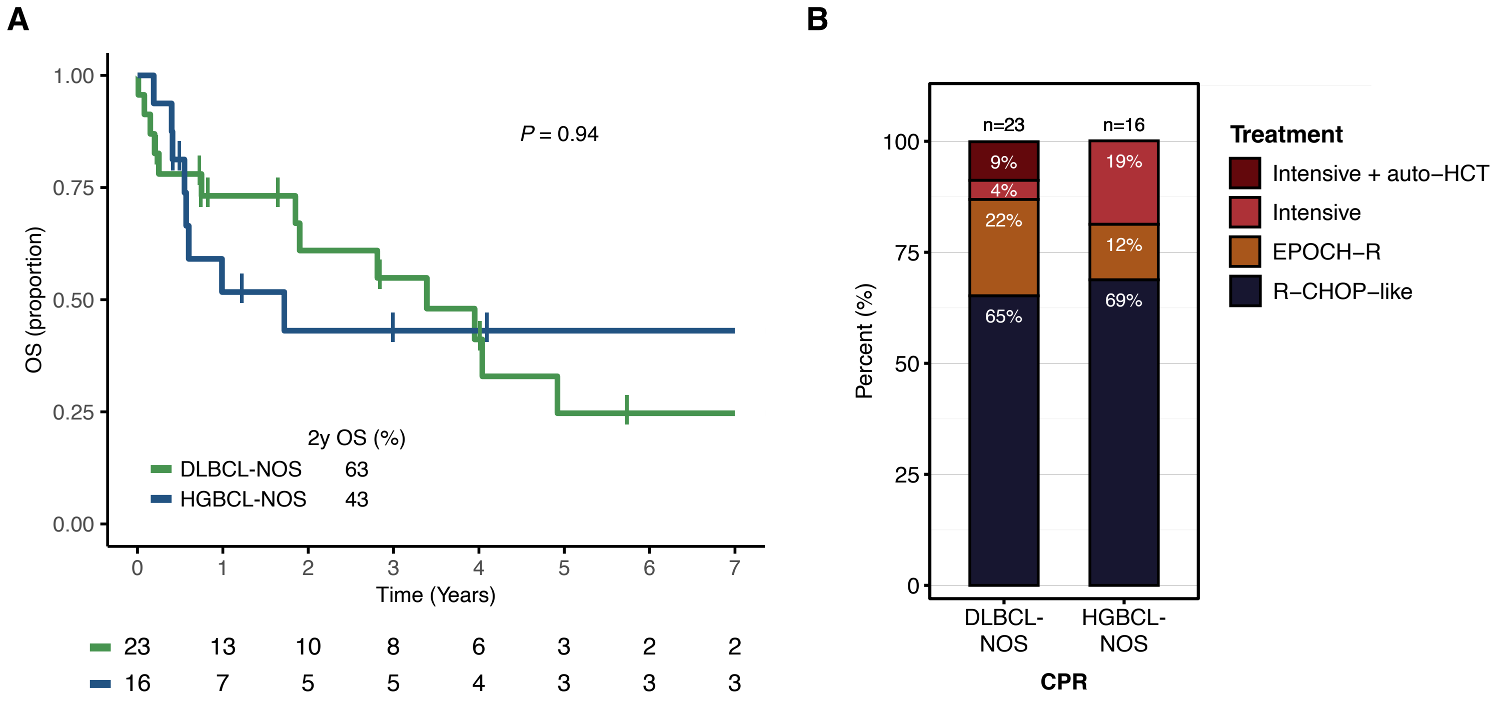


**Supplemental Figure 12. Outcomes in tumors confirmed as HGBCL-NOS or reclassified as DLBCL-NOS by CPR. (A)** Overall survival (OS) by CPR. OS was determined from the date of diagnosis to the date of last follow-up or death from any cause. **(B)** Treatment regimen stratified by CPR. P value was determined by the log-rank test.
